# ‘Necessity is the mother of invention’: Specialist palliative care service innovation and practice change in response to COVID-19. Results from a multi-national survey (CovPall)

**DOI:** 10.1101/2020.10.29.20215996

**Authors:** Lesley Dunleavy, Nancy Preston, Sabrina Bajwah, Andy Bradshaw, Rachel Cripps, Lorna K Fraser, Matthew Maddocks, Mevhibe Hocaoglu, Fliss EM Murtagh, Adejoke Oluyase, Katherine E Sleeman, Irene Higginson, Catherine Walshe, on behalf of the CovPall study team

## Abstract

**Background:** Specialist palliative care services have a key role in a whole system response to COVID-19. There is a need to understand service response to share good practice and prepare for future care.

**Aim:** To map and understand specialist palliative care services innovations and practice changes in response to COVID-19 (CovPall).

**Design:** Online survey of specialist palliative care providers, disseminated via key stakeholders. Data collected on service characteristics, innovations and changes in response to COVID-19. Statistical analysis included frequencies, proportions and means, and free-text comments were analysed using a qualitative framework approach.

**Setting/participants:** Inpatient palliative care units, home nursing services, hospital and home palliative care teams from any country.

**Results:** 458 respondents: 277 UK, 85 Europe (except UK), 95 World (except UK and Europe), 1 missing country. 54.8% provided care across 2+ settings; 47.4% hospital palliative care teams, 57% in-patient palliative care units, and 57% home palliative care teams. The crisis context meant services implemented rapid changes. Changes involved streamlining, extending and increasing outreach of services, using technology to facilitate communication, and implementing staff wellbeing innovations. Barriers included; fear and anxiety, duplication of effort, information overload, funding, and IT infrastructure issues. Enablers included; collaborative teamwork, pooling of staffing resources, staff flexibility, a pre-existing IT infrastructure and strong leadership.

**Conclusions:** Specialist palliative care services have been flexible, highly adaptive and have adopted a ‘frugal innovation’ model in response to COVID-19. In addition to financial support, greater collaboration is essential to minimise duplication of effort and optimise resource use.

**ISRCTN16561225:** https://doi.org/10.1186/ISRCTN16561225

**Key Statements:** *What is already known about the topic?:*

- Specialist palliative care is part of a whole healthcare system response to COVID-19.
- Services need to make practice changes in response to the global pandemic.

*What this paper adds:*

- Specialist palliative care services responded rapidly to COVID-19 in both planning for change and then adapting to needs and requirements.
- Services often relied on ‘improvisation’, ‘quick fixes’ and ‘making do’ when responding to the COVID-19 crisis.

*Implications for practice, theory or policy:*

- In addition to financial support, greater collaboration is essential to build organisational resilience and drive forward innovation, by minimising duplication of effort and optimising resource use.
- The effectiveness and sustainability of any changes made during the crisis needs further evaluation.

## Background

The COVID-19 pandemic is an example of a so-called ‘wicked problem’, constantly changing, difficult to define, and with multiple interdependencies^1^; what the army would call a VUCA situation: ‘volatile, uncertain, complex and ambiguous’^2^. Adaptations, flexibilities and innovative practices are necessary^3^. This includes innovative responses of healthcare systems, where hospice and palliative care services are an integral part of such a response to the COVID-19 pandemic.

Innovations are the tools used by organisations to influence or respond to environmental change and can encompass both radical and incremental innovation^4, 5^. The definition of innovation is discipline specific ^4^ and in health care it has been defined as; ‘a novel set of behaviours, routines, and ways of working that are discontinuous with previous practice, are directed at improving health outcomes, administrative efficiency, cost-effectiveness, or users’ experience and that are implemented by planned and coordinated actions’ (p582)^6^. A more ubiquitous definition, also used in health care^7, 8^, defines innovation more broadly as ‘an idea, practice or object that is perceived as new’ so a change in practice may be novel even if the same approach has been used elsewhere^9^ (p12).

Practice changes that may be regarded as innovations against these definitions are likely to be required and seen in response to COVID-19. Hospice and palliative services are argued to have an essential role^10^, especially with rapidly shifting priorities, new learning about the disease, potential shortages of drugs and equipment, and adjusting to workforce pressures and redeployments^11^. Commentaries indicate an initial rapidity of service changes with reports of new staffing and service delivery models, virtual care, and addressing shortages all in the context of infection control procedures and heightened fear and anxiety^12-16^. It is imperative that we consider which might be sustained as part of a ‘new normal’, and which may quietly fall away^17, 18^, with a focus on a learning mindset^19^. In this context, it is important that there is wide learning about how hospice and palliative care services have responded to the COVID-19 pandemic, so that effective innovations can be rapidly shared, and preparations made for future care, including second or third waves or other pandemic or emergency situations^20^.

## Methods

### Aim

To map and understand specialist palliative care services innovations and practice changes in response to COVID-19. This paper is part of the wider CovPall study that aims to understand the multi-national specialist palliative care response to COVID-19.

### Design

An online multi-national cross-sectional survey of hospice and specialist palliative care providers. This study is reported according to the STROBE^21^ and CHERRIES^22^ statements.

### Population and setting

Inpatient palliative care units, hospital palliative care teams, home palliative care teams and home nursing services were eligible to provide data, from any country.

### Sampling and recruitment

The aim of the recruitment strategy was to receive responses from all hospice and specialist palliative care services. Information was widely disseminated through key collaborators (e.g. Hospice UK, Marie Curie Care, Sue Ryder Foundation, European Association of Palliative Care), contacts through publicly available service directories, information provided on institutional websites, personal networks, and via the social media channels of investigators and key collaborators. Potential participants could contact the study team to receive a participant information sheet and link to complete the survey online. The clinical lead or their nominee completed the survey.

### Data collection

REDCap was used to securely build and host the online survey with closed and free text survey responses (see supplementary materials for the full survey). Sites could enter the data online directly (with a pause and return function), be sent the survey as a word document via email to complete and return electronically, or request to answer the survey questions via telephone or video conferencing with a member of the study team. Multiple questions with free-text response options within the survey addressed relevant areas for this analysis, and are presented in table 1.

**Table 1:**
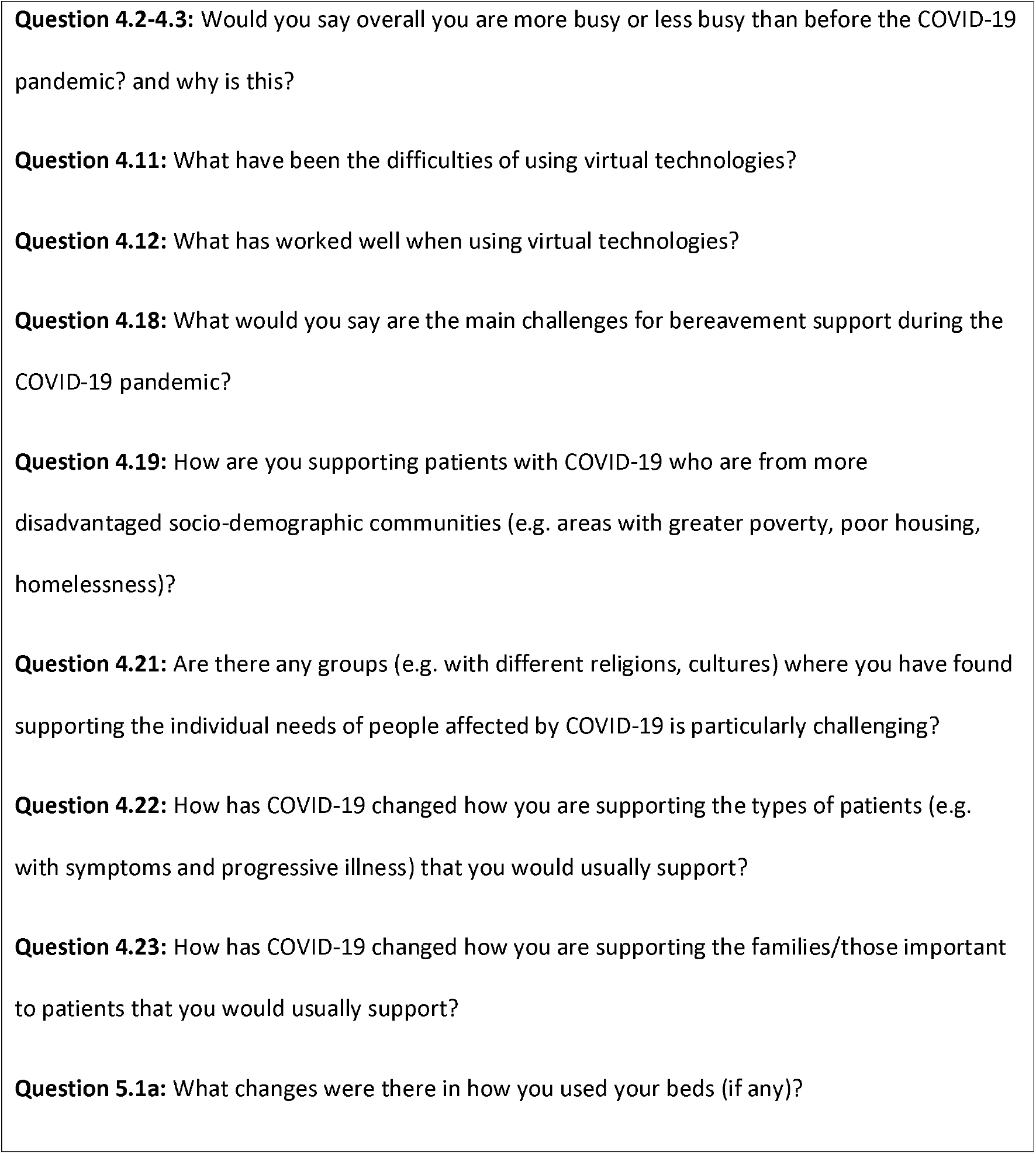
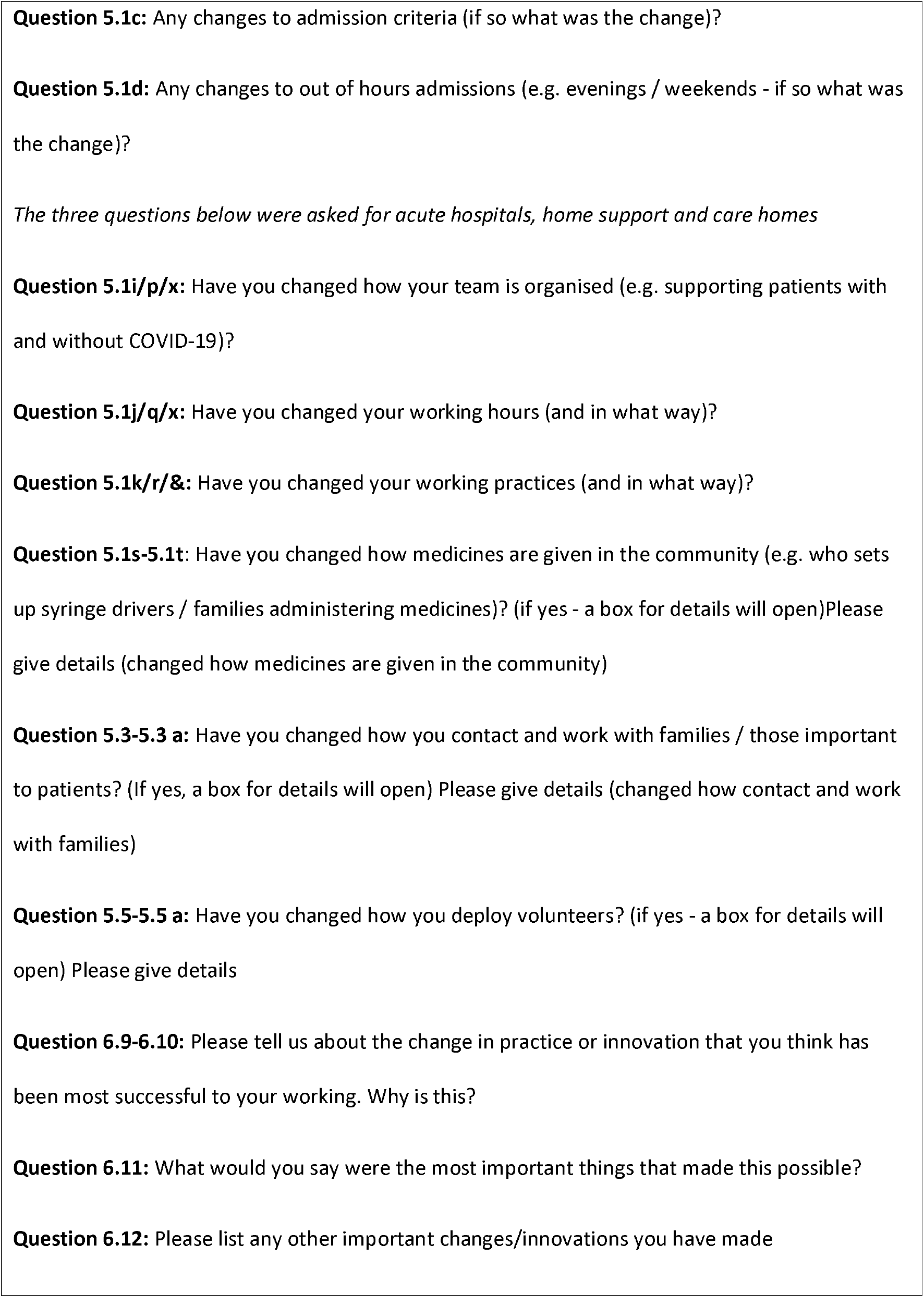
Free text response survey questions reported in this paper

### Data analysis

Anonymised data were exported to SPSS (for quantitative analysis using descriptive statistics, frequencies, proportions and means) and NVivo 12 (for analysis of free-text comments using a qualitative framework analysis approach)^23^. Continuous variables were expressed as means (SD) and medians (IQR) and categorical variables as counts and percentages. Missing data were not imputed. An analytical framework was initially developed by LD and CW through familiarisation with data, the framework was then applied to the free text data and refined, as appropriate, during the analysis process. LD and CW used analytical memos and charting to aid interpretation of the data.

### Research ethics and approvals

Research ethics committee approval for this study was obtained from King’s College London Research Ethics Committee (21/04/2020, Reference; LRS19/20-18541). The study was registered on the ISRCTN registry (27/07/2020, ISRCTN16561225).

#### Findings

The survey was open to responses from 23/04/2020 to 31/07/2020. Responses were received from 458 respondents: 277 UK, 85 Europe (except UK), 95 World (except UK and Europe), 1 missing country. The response rate could not be calculated as the survey denominator was unknown. Table 2 reports data on the characteristics of responding services and answers to the survey questions explored in the findings below.

**Table 2:** Characteristics of services responding to the survey and answers to the survey questions explored in the findings below.

|  | UK<br>(n = 277) | Europe (except UK)<br>(n = 85) | World (except Europe) (n = 95) |  |  | Total |
| --- | --- | --- | --- | --- | --- | --- |
|  |  |  | LIC/LMIC<br>(n = 17) | UMIC<br>(n = 19) | HIC<br>(n = 59) |  |
| **Role of respondents** (n/N, %)<sup>#</sup> |  |  |  |  |  |  |
| Medical director/ lead medical clinician | 97/274 (35.4%) | 42/85 (49.4%) | 11/17 (64.7%) | 7/19 (36.8%) | 26/58 (44.8%) | 183/453 (40.4%) |
| Nurse director/ lead nurse clinician | 69/274 (25.2%) | 8/85 (9.4%) | 2/17 (11.8%) | 2/19 (10.5%) | 7/58 (12.1%) | 88/453 (19.4%) |
| Other | 108/274 (39.4%) | 35/85 (41.2%) | 4/17 (23.5%) | 10/19 (52.6%) | 25/58 (43.1%) | 182/453 (40.2%) |
| Missing | 3 | - | - | - | 1 | 5<sup>+</sup> |
| **Type of service** (n/N, %) |  |  |  |  |  |  |
| Adult only | 247/277 (89.2%) | 68/85 (80%) | 7/17 (41.2%) | 9/19 (47.4%) | 40/59 (67.8%) | 371/458 (81%)* |
| Children only | 16/277 (5.8%) | 5/85 (5.9%) | 0/17 (0%) | 3/19 (15.8%) | 5/59 (8.5%) | 29/458 (6.3%)* |
| Both adult and children | 11/277 (4%) | 10/85 (11.8%) | 9/17 (52.9%) | 7/19 (36.8%) | 14/59 (23.7%) | 52/458 (11.4%)* |
| None indicated | 3/277 (1.1%) | 2/85 (2.4%) | 1/17 (5.9%) | 0/19 (0%) | 0/59 (0%) | 6/458 (1.3%)* |
| **Setting** (n/N, %) |  |  |  |  |  |  |
| Inpatient PC unit | 168/277 (60.6%) | 44/85 (51.8%) | 8/17 (47.1%) | 6/19 (31.6%) | 34/59 (57.6%) | 261/458 (57%)* |
| Hospital PC team | 135/277 (48.7%) | 26/85 (30.6%) | 9/17 (52.9%) | 10/19 (52.6%) | 37/59 (62.7%) | 217/458 (47.4%)* |
| Home PC team | 160/277 (57.8%) | 47/85 (55.3%) | 10/17 (58.8%) | 9/19 (47.4%) | 34/59 (57.6%) | 261/458 (57%)* |
| Home nursing | 92/277 (33.2%) | 15/85 (17.6%) | 1/17 (5.9%) | 4/19 (21.1%) | 7/59 (11.9%) | 119/458 (26%)* |
| Total | 277 | 85 | 17 | 19 | 59 | 458* |
| **Management type (n/N, %)\*** |  |  |  |  |  |  |
| Charitable / non-profit | 143/262 (54.6%) | 23/85 (27.1%) | 5/17 (29.4%) | 6/17 (35.3%) | 14/58 (24.1%) | 192/440 (43.6%)* |
| Public | 103/262 (39.3%) | 51/85 (60%) | 8/17 (47.1%) | 6/17 (35.3%) | 36/58 (62.1%) | 204/440 (46.4%)* |
| Private | 1/262 (0.4%) | 9/85 (10.6%) | 2/17 (11.8%) | 5/17 (29.4%) | 2/58 (3.4%) | 19/440 (4.3%)* |
| Other | 15/262 (5.7%) | 2/85 (2.4%) | 2/17 (11.8%) | 0/17 (0%) | 6/58 (10.3%) | 25/440 (5.7%)* |
| Missing | 15 | - | - | 2 | 1 | 18 |
| **Setting (n/N, %)** |  |  |  |  |  |  |
| None indicated | 3/277 (1.1%) | 1/85 (1.2%) | 1/17 (5.9%) | 0/19 (0%) | 0/59 (0%) | 5/458 (1.1%)* |
| Single setting | 110/277 (39.7%) | 49/85 (57.6%) | 8/17 (47.1%) | 11/19 (57.9%) | 24/59 (40.7%) | 202/458 (44.1%)* |
| Multiple settings (2 or more settings) | 164/277 (59.2%) | 35/85 (41.2%) | 8/17 (47.1) | 8/19 (42.1%) | 35/59 (59.3%) | 251/458 (54.8%)* |

| Information about services offered before COVID-19 pandemic |  |  |  |  |  |  |
| --- | --- | --- | --- | --- | --- | --- |
| **Use of remote consultations to help support patient care or education before COVID-19** |  |  |  |  |  |  |
| Telephone support for clinical care (n/N, %) | 179/277 (64.6%) | 54/85 (63.5%) | 13/17 (76.5%) | 11/19 (57.9%) | 45/59 (76.3%) | 303/458 (66.2%)* |
| **Use of remote consultations to help support patient care or education before COVID-19** |  |  |  |  |  |  |
| Telehealth/video support/e-learning for education (n/N, %) | 88/277 (31.8%) | 11/85 (12.9%) | 1/17 (5.9%) | 6/19 (31.6%) | 21/59 (35.6%) | 127/458 (27.7%)+ |
| **Use of remote consultations to help support patient care or education before COVID-19** |  |  |  |  |  |  |
| Telehealth/ video support/e-learning for clinical care (n/N, %) | 54/277 (19.5%) | 11/85 (12.9%) | 4/17 (23.5%) | 4/19 (21.1%) | 26/59 (44.1%) | 99/458 (21.6%)+ |
| Experience with suspected or confirmed COVID-19 |  |  |  |  |  |  |
| **Services with confirmed or suspected COVID-19 cases (n/N, %)<sup>#</sup>** | 248/264 (93.9%) | 60/83 (72.3%) | 9/17 (52.9%) | 7/17 (41.2%) | 33/58 (56.9%) | 358/440 (81.4%)* |
| Missing | 13 | 2 | - | 2 | 1 | 18 |
| **Services with staff with suspected or confirmed COVID-19 (n/N, %)<sup>#</sup>** | 238/262 (90.8%) | 55/83 (66.3%) | 4/16 (25%) | 7/17 (41.2%) | 30/58 (51.7%) | 335/437 (76.7%)* |
| Missing | 15 | 2 | 1 | 2 | 1 | 21 |
| **Service changes in response to COVID-19** |  |  |  |  |  |  |
| **Services who had changed in response to COVID-19 (n/N, %)<sup>#</sup>** | 255/263 (97%) | 74/82 (90.2%) | 14/17 (82.4%) | 15/17 (88.2%) | 53/57 (93%) | 412/437 (94.3%)* |
| Missing | 14 | 3 | - | 2 | 2 | 21 |
| **Changes in specific services** |  |  |  |  |  |  |
| **Services that reported changes in in-patient beds in their service (n/N, %)** | 117/255 (45.9%) | 25/74 (33.8%) | 4/14 (28.6%) | 4/15 (26.7%) | 20/53 (37.7%) | 170/412 (41.3%)* |
| **Changes in in-patient beds in your services (n/N, %)**<br>Number of beds |  |  |  |  |  |  |
| Increased | 41/117 (35%) | 5/28 (17.9%) | 1/4 (25%) | 0/4 (0%) | 1/21 (4.8%) | 48/174 (27.6%) |
| Stayed about the same | 34/117 (29.1%) | 10/28 (35.7%) | 2/4 (50%) | 4/4 (100%) | 7/21 (33.3%) | 57/174 (32.8%) |
| Decreased | 42/117 (35.9%) | 13/28 (46.4%) | 1/4 (25%) | 0/4 (0%) | 13/21 (61.9%) | 69/174 (39.7%) |
| **Services that reported changes in how they provide support for patients in their own homes (n/N, %)** | 152/255 (59.6%) | 32/74 (43.2%) | 8/14 (57.1%) | 6/15 (40%) | 30/53 (56.6%) | 228/412 (55.3%)* |
| **Number of patients needing support in their own homes (n/N, %)** |  |  |  |  |  |  |
| Increased | 60/152 (39.5%) | 15/32 (46.9%) | 1/8 (12.5%) | 4/6 (66.7%) | 13/31 (41.9%) | 93/229 (40.6%) |
| Stayed about the same | 72/152 (47.4%) | 11/32 (34.4%) | 4/8 (50%) | 1/6 (16.7%) | 15/31 (48.4%) | 103/229 (45%) |
| Decreased | 20/152 (13.2%) | 6/32 (18.8%) | 3/8 (37.5%) | 1/6 (16.7%) | 3/31 (9.7%) | 33/229 (14.4%) |
| **Services that reported changes in how medicines are given in the community (e.g. who sets up syringe drivers/families administering medicines (n/N, %)<sup>#</sup>** | 56/149 (37.6%) | 7/31 (22.6%) | 1/8 (12.5%) | 2/6 (33.3%) | 9/28 (32.1%) | 75/222 (33.8%) |
| **Services that reported changes in how they provide support for patients in care homes (including nursing homes) (n/N, %)** | 96/255 (37.6%) | 23/74 (31.1%) | 2/14 (14.3%) | 4/15 (26.7%) | 28/53 (52.8%) | 153/412 (37.1%)<sup>+</sup> |
| **Face to face contact with staff in care homes (including nursing homes) is (n/N, %)** |  |  |  |  |  |  |
| A lot more | 8/93 (8.6%) | 2/21 (9.5%) | 0/2 (0%) | 2/3 (66.7%) | 0/25 | 12/144 (8.3%) |
| Slightly more | 7/93 (7.5%) | 2/21 (9.5%) | 0/2 (0%) | 0/3 (0%) | 1/25 (4%) | 10/144 (6.9%) |
| About the same | 7/93 (7.5%) | 3/21 (14.3%) | 1/2 (50%) | 1/3 (33.3%) | 2/25 (8%) | 14/144 (9.7%) |
| Slightly less | 15/93 (16.1%) | 2/21 (9.5%) | 1/2 (50%) | 0/3 (0%) | 4/25 (16%) | 22/144 (15.3%) |
| Much less | 56/93 (60.2%) | 12/21 (57.1%) | 0/2 (0%) | 0/3 (0%) | 18/25 (72%) | 86/144 (59.7%) |
| **Services that reported use of virtual technologies (e.g zoom/teams) with** |  |  |  |  |  |  |
| **patients and families (n/N, %)** |  |  |  |  |  |  |
| A lot more | 149/252 (59.1%) | 25/71 (35.2%) | 6/13 (46.2%) | 6/15 (40%) | 33/53 (62.3%) | 219/405 (54.1%)<sup>+</sup> |
| Slightly more | 71/252 (28.2%) | 24/71 (33.8%) | 2/13 (15.4%) | 8/15 (53.3%) | 14/53 (26.4%) | 120/405(29.6%)* |
| About the same | 29/252 (11.5%) | 19/71 (26.8%) | 1/13 (7.7%) | 0/15 (0%) | 6/53 (11.3%) | 55/405(13.6%)<sup>+</sup> |
| Slightly less | - | - | - | - | - | - |
| Much less | 3/252 (1.2%) | 3/71 (4.2%) | 4/13 (30.8%) | 1/15 (6.7%) | 0/53 | 11/405 (2.7%)<sup>+</sup> |
| **Services that reported use of virtual technologies (e.g zoom/teams) with colleagues (n/N, %)** |  |  |  |  |  |  |
| A lot more | 236/254 (92.9%) | 46/72 (63.9%) | 10/14 (71.4%) | 8/15 (53.3%) | 45/53 (84.9%) | 345/409 (84.4%)<sup>+</sup> |
| Slightly more | 14/254 (5.5%) | 14/72 (19.4%) | 3/14 (21.4%) | 5/15 (33.3%) | 8/53 (15.1%) | 45/409 (11%)* |
| About the same | 2/254 (0.8%) | 10/72 (13.9%) | 0/14 | 2/15 (13.3%) | 0/53 | 14/409 (3.4%)<sup>+</sup> |
| Slightly less | 1/254 (0.4%) | 2/72 (2.8%) | 0/14 | 0/15 | 0/53 | 3/409 (0.7%)<sup>+</sup> |
| Much less | 1/254 (0.4%) | 0/72 | 1/14 (7.1%) | 0/15 | 0/53 | 2/409 (0.5%)<sup>+</sup> |
| **Bereavement support** |  |  |  |  |  |  |
| **Bereavement support (n/N, %)** |  |  |  |  |  |  |
| A lot more | 32/193 (16.6%) | 2/53 (3.8%) | 1/13 (7.7%) | 0/12 | 2/41 (4.9%) | 37/313 (11.8%)<sup>+</sup> |
| Slightly more | 49/193 (25.4%) | 14/53 (26.4%) | 1/13 (7.7%) | 5/12 (41.7%) | 9/41 (22%) | 78/313 (24.9%)<sup>+</sup> |
| About the same | 76/193 (39.4%) | 24/53 (45.3%) | 5/13 (38.5%) | 2/12 (16.7%) | 21/41 (51.2%) | 129/313 (41.2%)* |
| Slightly less | 23/193 (11.9%) | 9/53 (17%) | 1/13 (7.7%) | 4/12 (33.3%) | 6/41 (14.6%) | 43/313 (13.7%)+ |
| Much less | 13/193 (6.7%) | 4/53 (7.5%) | 5/13 (38.5%) | 1/12 (8.3%) | 3/41 (7.3%) | 26/313 (8.3%)+ |

Note: UK = United Kingdom, LIC = Low Income Countries, LMIC = Lower Middle Income Countries, UMIC = Upper Middle Income Countries, HIC = High
Income countries, PC = Palliative Care

The overarching categories identified in the analysis included ‘the crisis context’, the changes made (streamlining access, extending services, increasing outreach, using communication technology and implementing innovations for staff wellbeing) and the enablers and barriers for change (see figure 1). Exemplar data extracts for each category and subcategory are presented in table 3 and in the narrative below.

**Table 3:**
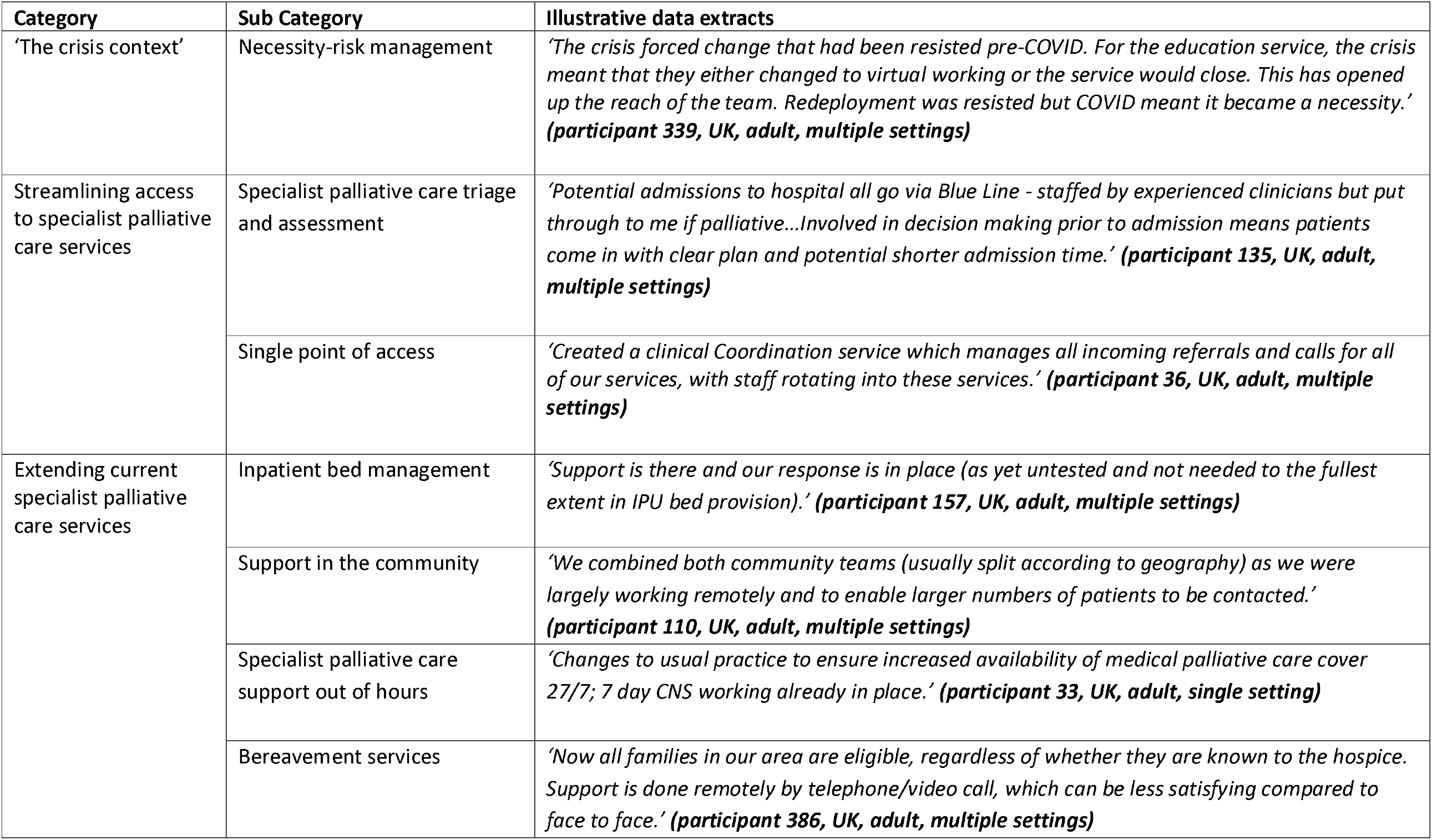
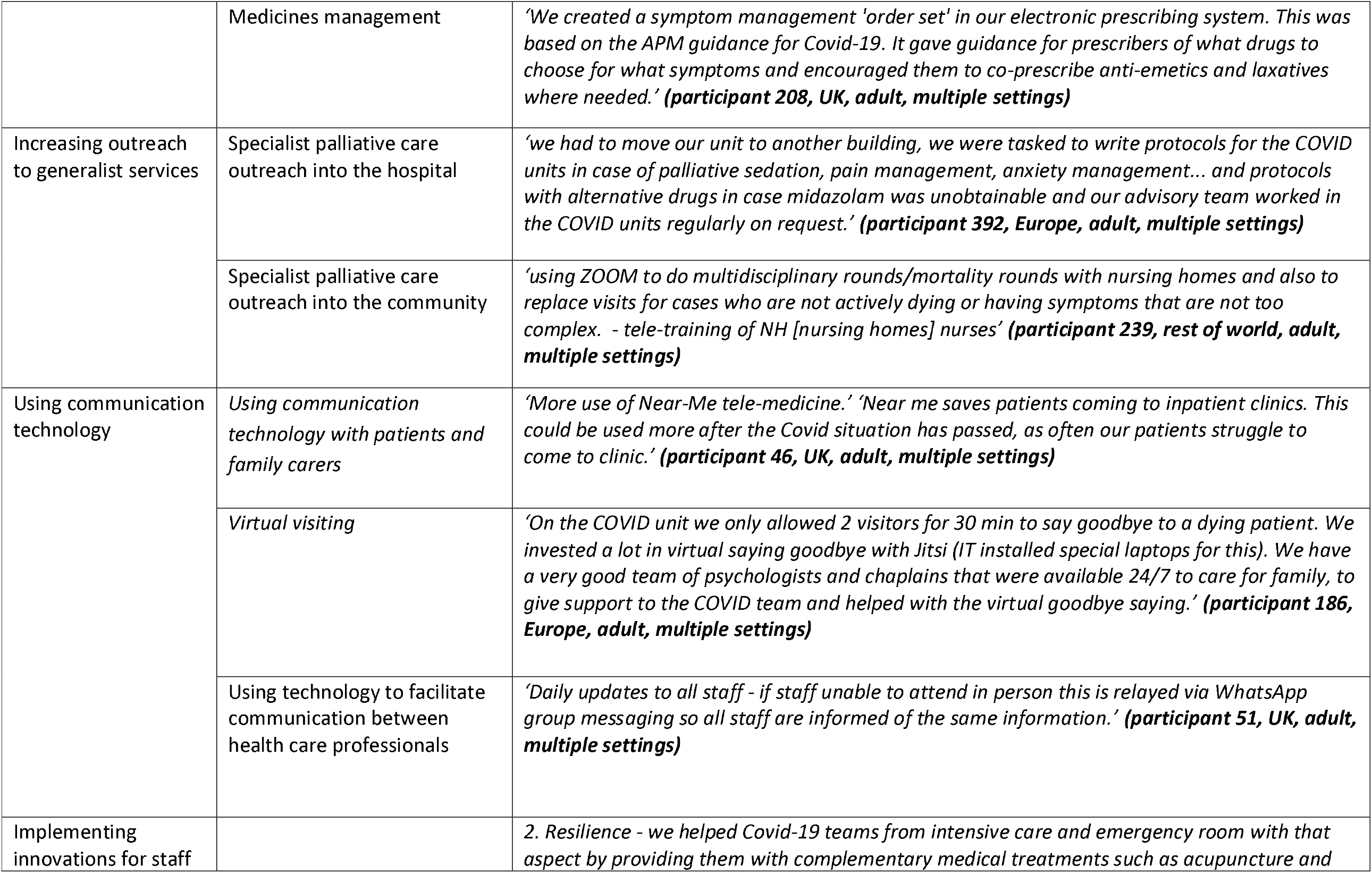
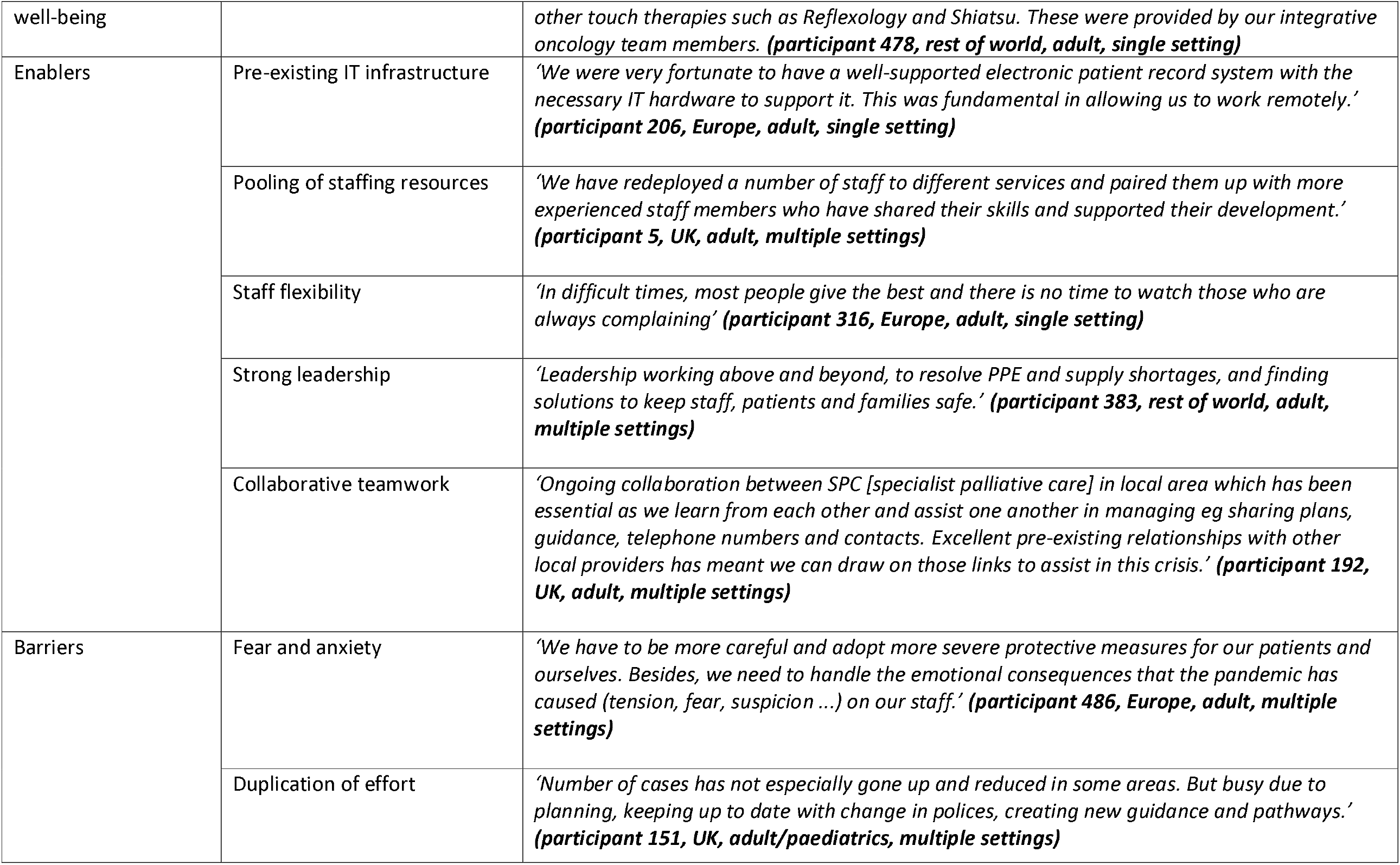
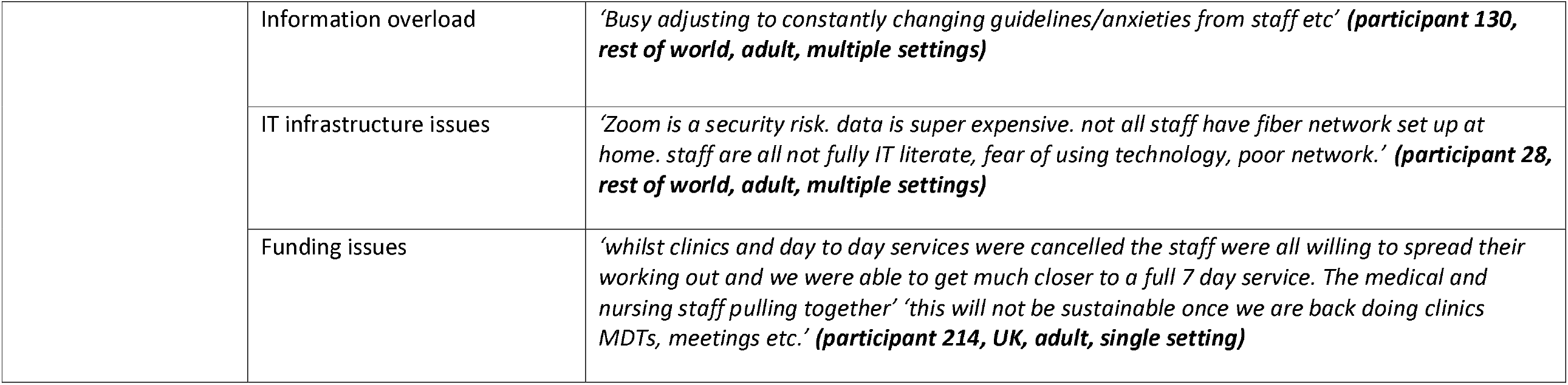
Categories and sub-categories

**Figure 1:**
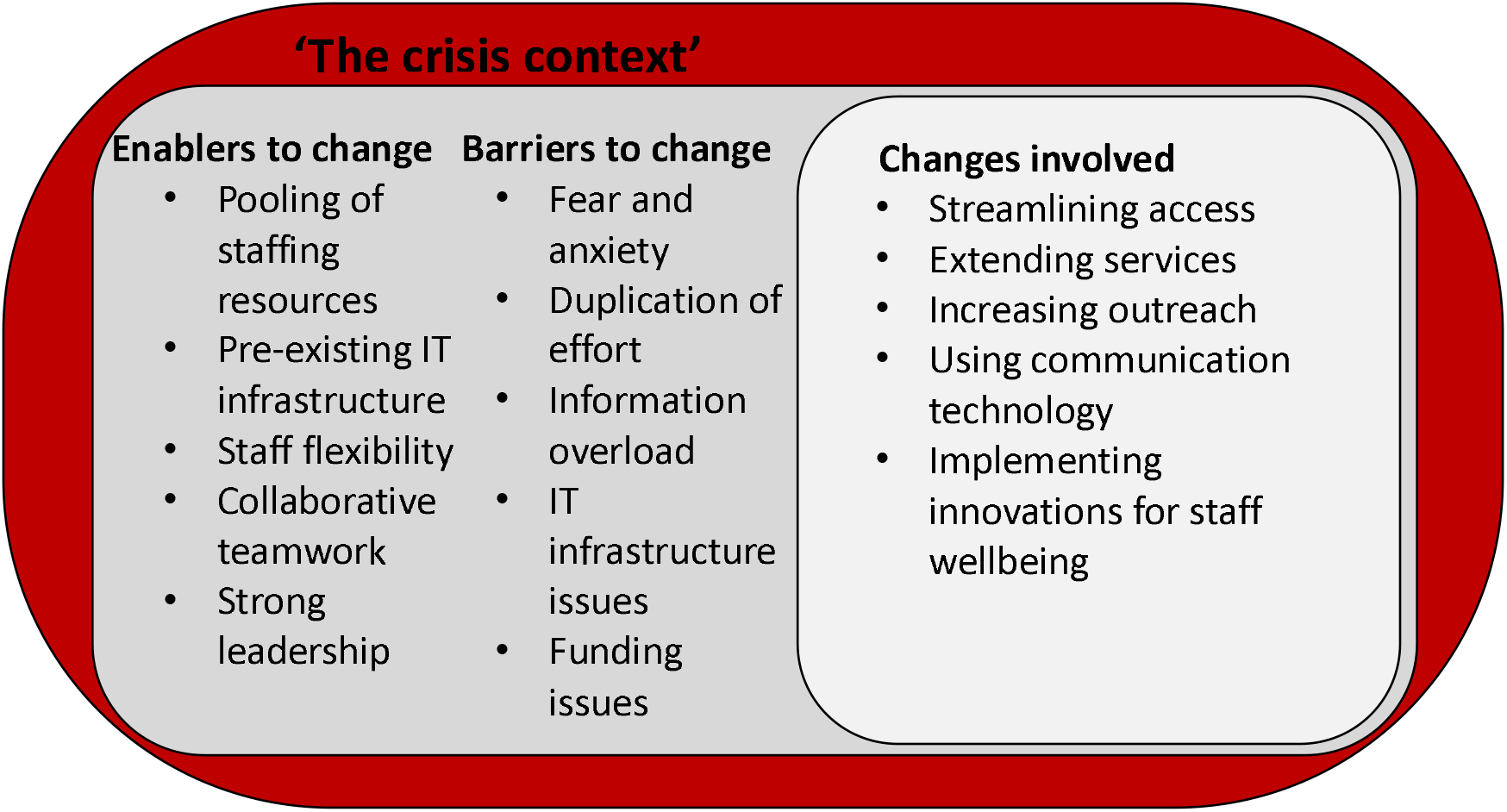
Overarching analytical categories

#### The crisis context

All services had to change, often rapidly, to prepare for, and respond to, the anticipated and actual impact of the COVID-19 pandemic. Services often initiated changes that had been previously considered but rejected or resisted.

#### Streamlining access to specialist palliative care services

Specialist palliative care triage and assessment/single point of access The pandemic led to changes in how referrals to services were received, assessed and managed, both initially and on an ongoing basis. This included, for example, proactively seeking referrals, loosening or tightening referral criteria, such as not accepting patients for respite care, and the use of telephone advice lines:

> *‘Setting up a regional single point of access with two other local hospices and pooling resources so patients, families and HCPs [health care professionals] have one number to call to gain advice, support or rapid discharge, admission to a hospice etc*.*’ ‘Something we have been working towards for a long time, but very slowly. The pandemic has allowed us to set it up in 10 days and iron out problems as we’ve gone. People have been a lot more collaborative*.*’ (****participant 92, UK, adult, multiple settings)***

How referrals for people without COVID-19 and with confirmed or suspected COVID-19 were dealt with was based on both clinical need and how to manage the risk of infection for patients, family carers and staff:

> *‘Triage telephone assessment prior to visits and in maintaining on going symptom management support*.*’ ‘It has enabled scrutiny of issues to prioritise a visit*.*’* ***(participant 47, UK, paediatric, multiple settings)***

Telephone calls were used to keep in regular contact with patients and family carers including proactive calls for patients who were stable but isolated:

> *‘Calls to all patients to gauge vulnerability, how they would get meds/ shopping in lockdown, what their supports were and specific form for this completed to help us know who to focus on-these patients received extra connection calls and support*.*’* ***(participant 421, rest of world, adult, multiple settings)***

Ongoing telephone support would also be provided for those patients who were unable to attend outpatients or day therapy services because of restrictions. Services wanted to reassure patients and family carers that ‘*the care is still here, it just looks different*’ ***(participant 42, UK, adult, single setting)***.

#### Extending current specialist palliative care services

##### Inpatient bed management

41.3 % of services who had changed in response to COVID-19 reported changes in inpatient beds in their service with 27.6 % reporting an increase in bed numbers. This was often in situations where dedicated beds previously did not exist, or else increasing capacity by using space in flexible ways:

> ‘*Having palliative care beds in the hospital, supported by palliative care doctors’ ‘early lobbying for a palliative care ward from palliative care team*.*’* ***(participant 61, UK, adult, single setting)***
>
> *‘We re-configured our well-being centre to provide 12 extra IPU [In-patient unit] beds. We have stopped providing respite during the pandemic to open up beds. We are admitting routinely 7 days per week*.*’* ***(participant 94, UK, adult, multiple setting)***

When reconfiguring inpatient areas, services needed to take account of ‘hot’ areas or zones where those with suspected or confirmed COVID-19 were cared for and ‘cold areas’ where people not suspected of having COVID-19 were cared for:

> *‘In patient area in acute hospital repurposed as COVID cohort ward for end of life care; second ward in community hospital previously used as ‘hospice’ reassigned as ‘clean’ palliative care area*.*’* ***(participant 7, UK, adult, multiple settings)***

Some also reported that inpatient bed numbers stayed the same, decreased and that additional beds were not always needed.

##### Support in the community

55.3 % of services that had changed due to COVID-19 said there had been changes in how they provided support for patients in their own home with 40.6 % saying the number of patients needing support at home had increased. There was a shift in patient need from the inpatient to the community setting in some areas:

> *‘We have amalgamated a number of different teams i*.*e. CNS [Clinical Nurse Specialist], H@H [Hospice at Home], AHP [Allied Health Professionals], IPU [Inpatient Unit] into 3 community locality multi-professional teams. These provide both specialist care and in addition, personal care for anyone in the last 3 months of life. This means that any face to face visiting might occur for instance between a CNS [Clinical Nurse Specialist] and a HCA [Health Care Assistant] so that both forms of specialist and personal care can be given. This reduces footfall within patient’s homes. It also helps with staffing*.*’* ***(participant 235, UK, adult, multiple settings)***

##### Specialist palliative care support out of hours

Some respondents described extending their current out of hours medical and/or nursing provision during the peak of the pandemic to support generalist palliative care providers and facilitate hospice inpatient admissions out of hours:

> *‘We moved to a 7 day onsite service to be able to see patients that were referred at weekends and who had significant symptoms deteriorating quickly. This also allowed up to check in with areas of the hospital who were dealing with larger number of deaths than normal and try support patients and staff early were possible*.*’* ***(participant 351, UK, adult, single setting)***

Medical and nursing shift patterns were sometimes adjusted to accommodate the increase in out of hours provision. Some services reported that routine hours of working had resumed in their locality.

##### Bereavement services

36.7 % of respondents reported that they were providing slightly more or a lot more bereavement support than before the pandemic with 41.2 % offering about the same. Some were offering support to those not directly under the care of the specialist palliative care service, including to nursing home staff, and educating others on how to provide bereavement care:

> *‘reorganising the family support team to work virtually and provide more education and support to wider groups outside of the hospice. e*.*g support for staff outside of the hospice on how to support someone with bereavement*.*’* ***(participant 194, UK, adult, multiple settings)***

Services were starting to see referral numbers pick up and were planning for the anticipated increase in those requiring bereavement support as a result of the pandemic.

##### Medicines management

The pandemic led to changes in how medicines were managed with routine practices and processes adapted to reduce infection risk and unnecessary visits, such as single use syringe drivers and specialist palliative care professionals administering medication in services where this was not norm. New processes were set up to improve access to symptom control medication:

> *‘Palliative care team now administering medications in the home where previously this was only done by district nurses. Grab Bags available for prescribers to take out medications to the house for administration (not to be left in the house). Now have PGDs (Patient Group Directives) that enable non-prescribers to take anticipatory medications out on visits in a Grab Bag and administer up to 3 PRN doses of Morphine/Midazolam/Buscopan/ /Levomepromazine under specific circumstances*.*’* ***(participant 367, UK, adult, multiple settings)***

33.8 % of services who had changed how they provide support for patients in their homes said they changed how medicines were given in the community. In those areas where policies were not already in place, policies were developed to support carer administration of subcutaneous medication and/or routes of administration were changed to oral.

#### Increasing outreach to generalist palliative care services

##### Specialist palliative care ‘outreach’ into the hospital

Hospital specialist palliative care teams often shifted from a responsive to a proactive model of care as patients with COVID-19 could deteriorate and die rapidly and some of those providing direct care lacked end of life care experience. Teams proactively engaged with clinicians in areas where COVID-19 patients were being cared such as intensive care units, emergency departments and respiratory wards. They focused on; developing and disseminating COVID-19 symptom control guidelines, providing symptom control advice, supporting colleagues with complex treatment escalation or withdrawal decisions, visiting patients as necessary, and providing end of life care training and support including how to communicate with relatives over the telephone. Training and guidelines needed to be brief and rapidly developed:

> *‘Staff on wards caring for patients with coronavirus identified that the biggest challenge was communicating with relatives over the telephone. Our hospital specialist palliative care team, led by the registrar, developed one-page guides to support this’* ***(participant 31, UK, Adult, multiple settings)***

##### Specialist palliative care ‘outreach’ into the community

59.7 % of services who had changed how they provided support for patients in care homes (including nursing homes) reported that their face-to-face contact with care home staff was much less during this time. Prior to COVID-19 only 27.7% of respondents provided telehealth/video support/e-learning for education. Specialist palliative care services proactively contacted care homes to offer support and advice and education was also provided from a distance:

> *‘They [medical staff] have set up twice weekly webinars for the wider health community, including GPs, nurses and nursing homes as well as a new advice email for GPs*.*’* ***(participant 330, UK, Adult, multiple settings)***

#### Using communication technology

Services were forced to adopt the use of technology so that some clinical services could continue to operate. Patients could decline inpatient admission or face-to-face visits as they were fearful of contracting the virus and were concerned about the visiting restrictions. Some clinical staff, including those who were shielding, worked from home when able, to maintain social distancing.

##### Using communication technology with patients and family carers

Prior to COVID-19, only 21.6% of services used telehealth/video support/e-learning for clinical care with 83.7 % of services that changed due to COVID-19 reporting that they were using virtual technologies with patients and families a lot more or slightly more during the pandemic. Generic digital platforms were used for communication such as Zoom, Skype, WhatsApp and Facebook. Hospice day therapy services were also provided off site using this technology such as complementary therapies via Skype and ‘Time to create’ via a Hospice YouTube channel. There were also reports of telemedicine being used, electronic care plans and applications to facilitate symptom assessment, virtual ward rounds and admission assessments:

> *‘…our physician has been making virtual rounds which have been very successful through the use of the app*.*’ ‘We were well set up to use technology for reporting patient care, as well as general updates. During the COVID pandemic, this allowed us to maximize the use of this app to extend it to allowing for virtual rounds with the doctor, as well as to complete virtual intake assessments with referrers*.*’* ***(participant 218, rest of world, adult, single setting)***

Using digital technology could be challenging especially with those who were hard of hearing, very sick or not previously known to the team. Not everyone had access to a computer, smart phone or the internet and connectivity could be an issue in rural areas. Some patients, particularly older people, were not keen to engage with digital technology. Prior to COVID-19, 66.2 % of services already used telephone support for clinical care and respondents reported that its use increased during the pandemic:

> *‘There has been more telephone contact with our usual community patients, either due to our choice or the patient’s. This has been ok but not ideal especially for difficult conversations or for new patients. It has however been more time efficient*.*’* ***(participant 19, UK, adult, multiple settings)***

Lack of closeness and human contact were reported as issues with remote working. Volunteer befriending, bereavement support and hospice day therapy services were also provided by telephone.

##### Virtual visiting

The pandemic led to visiting restrictions being imposed within inpatient units which caused distress to patients, family carers and staff members. Services utilised technology to facilitate ‘virtual visits’ and ensure lines of communication between patients and carers were kept open:

> *‘The iPads which we managed to raise through charity donations - through the use of our Face Book page - are now available on all wards with the support of our IT team and IG teams have allowed many families to speak or even just see their loved ones*.*’* ***(participant 175, UK, adult, single setting)***

A less costly and simpler strategy was the use of postcards for e-mail and telephone messages from relatives to be given or read out to patients.

##### Using technology to facilitate communication between health care professionals

84.4 % of services that changed in response to COVID-19 reported that they were using virtual technologies (e.g. zoom/teams etc.) with colleagues a lot more than before the pandemic. It was used to facilitate communication within specialist palliative care teams, across specialist palliative care services and with generalist clinicians and external partner organisations. Respondents felt the benefits included increased efficiency by reducing travel time, keeping the team connected and up to date with the ever-changing situation as well as helping to facilitate the support process:

> *‘firstly the support we gave our teams on a daily level. we realized they were leaving their family at home to visit patients and we perceived that as a vulnerable situation to many team members. we compiled a extensive reaching out plan to all the teams on an individual, sectorial, regional and nationwide level with phone support and zooms by professional management and general management*.*’* ***(participant 247, rest of world, adult/paediatric, multiple settings)***

Using technology required an understanding of virtual meeting ‘etiquette’ and could be exhausting with one respondent describing it as ‘Zoom fatigue’ ***(participant 222, rest of world, adult/paediatric, single setting)***.

#### Implementing innovations for staff well being

76.7 % of services had staff with suspected or confirmed COVID-19 so services needed to manage the impact of COVID-19 on staff wellbeing:

> *‘It has been awful to witness the sorrow and pain of some families, when they have been separated in times of crisis*.*’* ***(participant 225, Europe, adult, single setting)***

Different strategies were implemented to promote staff well-being while ensuring social distancing. The importance of checking in with staff on a regular basis, including those furloughed, to keep them informed of the ever-changing situation, and provide opportunities for staff debriefing was reported. More practical strategies included; free car parking and meals, help with child care, and virtual yoga or complementary therapy:

> *‘Our hospice clinic room has been repurposed as a staff room - ‘the bubble’ - again this is something we’d been talking about for several months but had been hard to agree on where it would be; in the event two colleagues did a makeover of the room one day, including soft furnishings and hand creams / handmade scrubs bags, handmade fixtures for masks to avoid skin irritation, chocolate etc - feels important as one way to show colleagues are valued*.*’* ***(participant 128, UK, adult, multiple settings)***

#### Enablers and barriers to change

Changes in practice occurred in a crisis context and in some instances this accelerated changes that had been previously planned or hoped for:

> *‘Necessity is the mother of invention. The situation has forced us to be creative and some of the ways we have done this will stay with us post covid*.*’* ***(participant 428, UK, adult, multiple settings)***.
>
> *‘Streamlining of two services managed by different organisations (acute trust and hospice) which we have wanted to achieve for decades!’ ‘Appetite for change on both sides to work together and disregard former barriers to put the interests of patients first - long may it last!’* ***(participant 388, UK, adult, multiple settings)***

Respondents identified several factors that they felt enabled the imposed changes to be implemented swiftly into clinical practice. These included; pooling of staffing resources, staff flexibility, strong leadership, collaborative teamwork and having a pre-existing IT infrastructure:

> *‘The community SPC already used a virtual delegating system for all visits, the eShift system…*.*This has been expanded during the pandemic to include all telephone consultations and reviews. This practice has allowed the team to convert to telephone consultations in a streamline manner already using the eShift structure. It has also allowed clinicians to work from home whilst still being supervised and supported clinically*.*’* ***(participant 173, UK, adult, multiple settings)***

A lack of access to basic IT equipment such as cameras, microphones or laptops, poor Wi-Fi or internet connection and there being too many digital platforms was a barrier to change. The need to implement remote working rapidly meant there was no time for training and staff could lack confidence and be unfamiliar with the technology but needed to learn quickly. Emergency COVID-19 funding was available but the sustainability of out of hours services without adequate funding was raised as an issue:

> *‘7 day service as patients and families had equitable services 7 days a week. Staff felt supported and wanted this sustained. Business case for 7 day services escalated*.*’* ***(participant 346, UK, adult, single setting)***

Changes were also being implemented at a time of heightened anxiety and fear and when services needed to handle and digest ever changing information which one respondent described as an *‘infodemic’* ***(participant 107, UK, adult, multiple settings)***. There was also evidence of effort duplication with services producing their own guidelines, procedures and policies.

## Discussion

### Main findings

Hospice and specialist palliative care services had to implement changes rapidly to respond to the anticipated and actual impact of the COVID-19 pandemic. Changes in practice involved streamlining, extending and increasing outreach of services, as well the use of communication technology and innovations for staff wellbeing. A number of barriers and enablers to change were evident such as fear and anxiety, duplication of effort, pooling of staffing resources and collaborative teamwork.

### What this study adds

Changes seen do not reflect the standard literature on the diffusion of innovations^6, 9^. Standard forms of innovation require planning and funding, often impossible when responding to an unforeseen event like the COVID-19 pandemic ^24, 25^. The term improvisation rather than innovation has been used in crisis management ^24-27^, as organisations are required to be creative by using, adjusting and recombining existing resources, structures and processes to manage the impact of a crisis ^28^. In these circumstances, resistance to change is limited as there is an acceptance that ‘normal’ rules no longer apply and a collective identity develops, as seen in this study, with clinicians no longer working in professional silos and previously resisted technology being used ^25^. Whilst used in a different context, such limited resistance to change resonates with Klein’s ^29^ concept of the ‘shock doctrine’ in which extreme crises (such as COVID-19) pertain the power to ‘shock’ systems and, in doing so, shake up socio-cultural norms to the extent that new changes – that may have been previously resisted - can be made quicker and easier than usual.

In this study, services had to rely on a ‘quick fix’, ‘making do’, being flexible and thinking in a frugal way. So called ‘frugal’ or ‘Jugaad’ innovation can challenge standard definitions of innovation^30^. The aim is to provide low cost solutions to problems in environments that have resource constraints ^31, 32^, and has been used in health care in economically disadvantaged communities ^33^, including in the context of palliative care ^34^.

Specialist palliative care services demonstrated considerable flexibility and ‘frugal’ innovation, and will continue to play an important role in managing the impact of the COVID-19 pandemic^10, 35^. Organisations need to build flexible and resilient systems so they can be responsive to the ongoing crisis, including threats to their income as a result of an economic downturn, and any future increases in infection rates. Both national and international collaboration, and coordinated action is required to optimise resource use and avoid duplication of effort, particularly in relation to training and guideline development, while maintaining high standards of care. This need for greater collaboration was highlighted in a recent review that found a dearth of comprehensive international COVID-19 guidance on palliative care for nursing homes ^36^.

### Strengths and weaknesses of the study

This study is a large multi-national survey of specialist palliative care services response to the COVID-19 pandemic. Free text responses provided useful insights into how and why services made changes to their routine ways of working in response to the crisis. The survey was completed by service leads so the findings may present an overly positive view of the changes made and may not reflect the views of other practitioners working within the services taking part in this study. Negative aspects of the changes made may also not have been captured due to the wording of questions in the survey. More detailed survey responses were also generally provided by those who were native English speakers.

Data were collected at a single time point so how useful and sustainable the changes were has not been captured. A successful frugal innovation or improvisation may be retained but may not be useful unless there is a similar future crisis ^27^. Changes in practice may lead to unethical practices and negative outcomes as resource scarcity may, in some instances, simply undermine the quality of care ^25^. The challenge of implementing remote clinical consultations rapidly during the pandemic with limited resources, for example, has been raised ^37^ and how sustainable changes are beyond the pandemic without the necessary infrastructure being in place has been questioned ^38^. This issue is particularly pertinent to palliative care where funding for services in some countries relies heavily on charitable funding. Further qualitative case study research is planned to explore in greater depth how services responded to the pandemic and why they did or did not implement particular changes into practice and whether changes were sustained and viewed as effective.

### Conclusion

Specialist palliative care services have responded rapidly to the COVID-19 pandemic. Services have demonstrated considerable flexibility and relied on ‘frugal innovation’ when responding to the crisis. Enablers to change included collaborative teamwork, pooling of staffing resources, staff flexibility, a pre-existing IT infrastructure and strong leadership. In addition to financial support, greater collaboration is essential to build organisational resilience by minimising duplication of effort and resource use. The effectiveness of any changes made during the crisis needs continued evaluation.

## The CovPall study group

### CovPall Study Team

Professor Irene J Higginson (Chief Investigator), Dr Sabrina Bajwah (Co-I), Dr Matthew Maddocks (Co-I), Professor Fliss Murtagh (Co-I), Professor Nancy Preston (Co-I), Dr Katherine E Sleeman (Co-I), Professor Catherine Walshe (Co-I), Professor Lorna K Fraser (Co-I), Dr Mevhibe B Hocaoglu (Co-I), Dr Adejoke Oluyase (Co-I), Dr Andrew Bradshaw, Lesley Dunleavy and Rachel L Cripps.

### CovPall Study Partners

Hospice UK, Marie Curie, Sue Ryder, Palliative Outcome Scale Team, European Association of Palliative Care (EAPC), Together for Short Lives and Scottish Partnership for Palliative Care.

## Data Availability

Applications for use of the survey data can be made for up to 10 years, and will be considered on a case by case basis on receipt of a methodological sound proposal to achieve aims in line with the original protocol. The study protocol is available on request. All requests for data access should be addressed to the Chief Investigator via the details on the CovPall website (https://www.kcl.ac.uk/cicelysaunders/research/evaluating/covpall-study, and) and will be reviewed by the Study Steering Group.

## Declarations

### Authorship

IJH is the grant holder and chief investigator; KES, MM, FEM, CW, NP, LKF, SB, MBH and AO are co-applicants for funding. IJH and CW with critical input from all authors wrote the protocol for the CovPall study. MBH, AO, RC and LD co-ordinated data collection and liaised with centres, with input from IJH, FEM, CW, NP and LKF. LD, CW, AO, MBH, IJH and LKF analysed the data. All authors had access to all study data, discussed the interpretation of findings and take responsibility for data integrity and analysis. LD and CW drafted the manuscript. All authors contributed to the analysis plan and provided critical revision of the manuscript for important intellectual content. IJH is the guarantor.

### Ethics and consent

Research ethics committee approval for this study was obtained from King’s College London Research Ethics Committee (21/04/2020, Reference; LRS19/20-18541). ISRCTN16561225. Completion of survey indicated the participant had consented to the study.

## Acknowledgements

This study was part of CovPall, a multi-national study, supported by the Medical Research Council, National Institute for Health Research Applied Research Collaboration South London and Cicely Saunders International. We thank all collaborators and advisors. We thank all participants, partners, PPI members and our Study Steering Group. We gratefully acknowledge technical assistance from the Precision Health Informatics Data Lab group (https://phidatalab.org) at National Institute for Health Research (NIHR) Biomedical Research Centre at South London and Maudsley NHS Foundation Trust and King’s College London for the use of REDCap for data capture. Sites who contributed to this work can be found in supplementary file 3.

