## Supplementary figures and images for "‘Necessity is the mother of invention’: Specialist palliative care service innovation and practice change in response to COVID-19. Results from a multi-national survey (CovPall)"

### Supplementary file 1

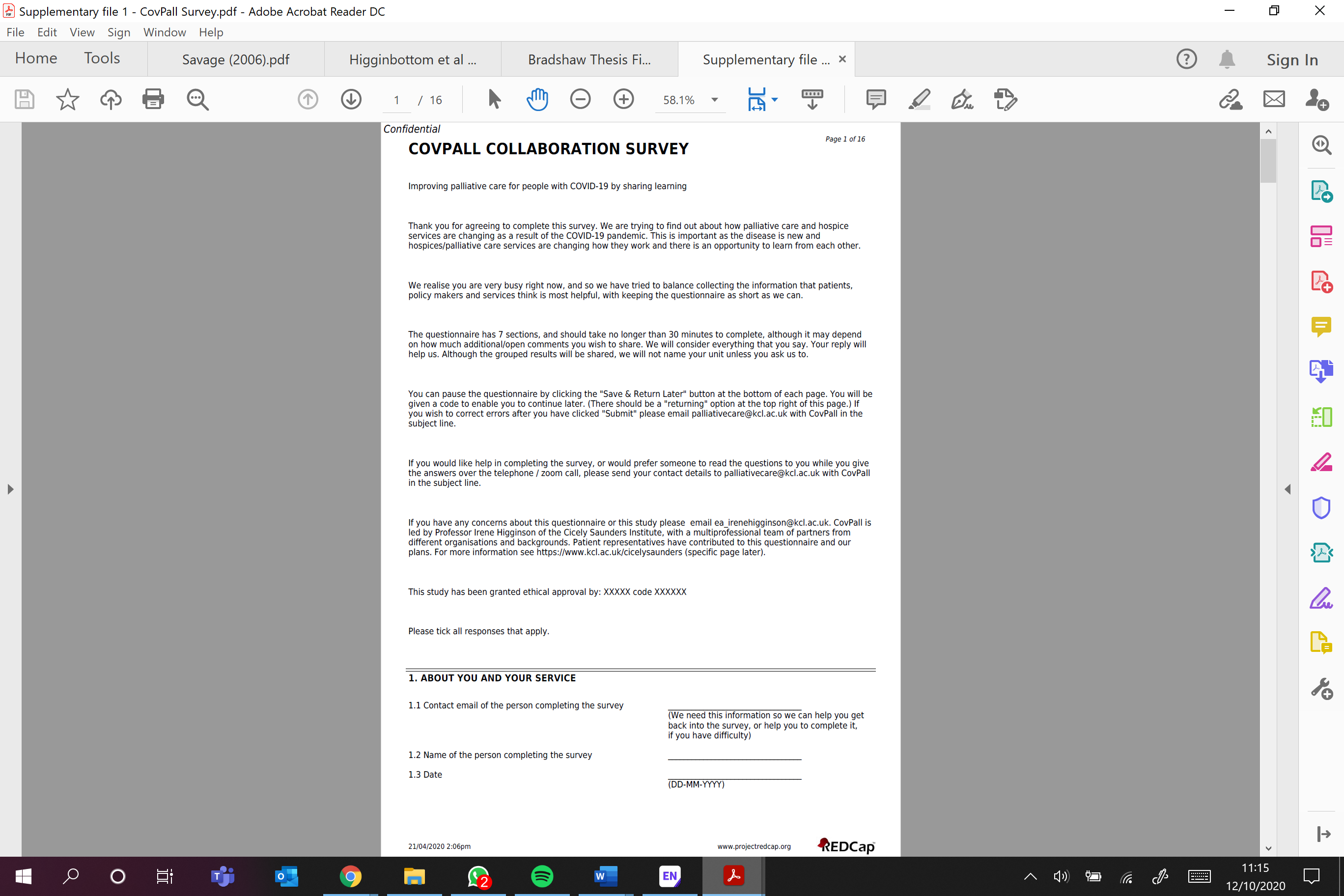

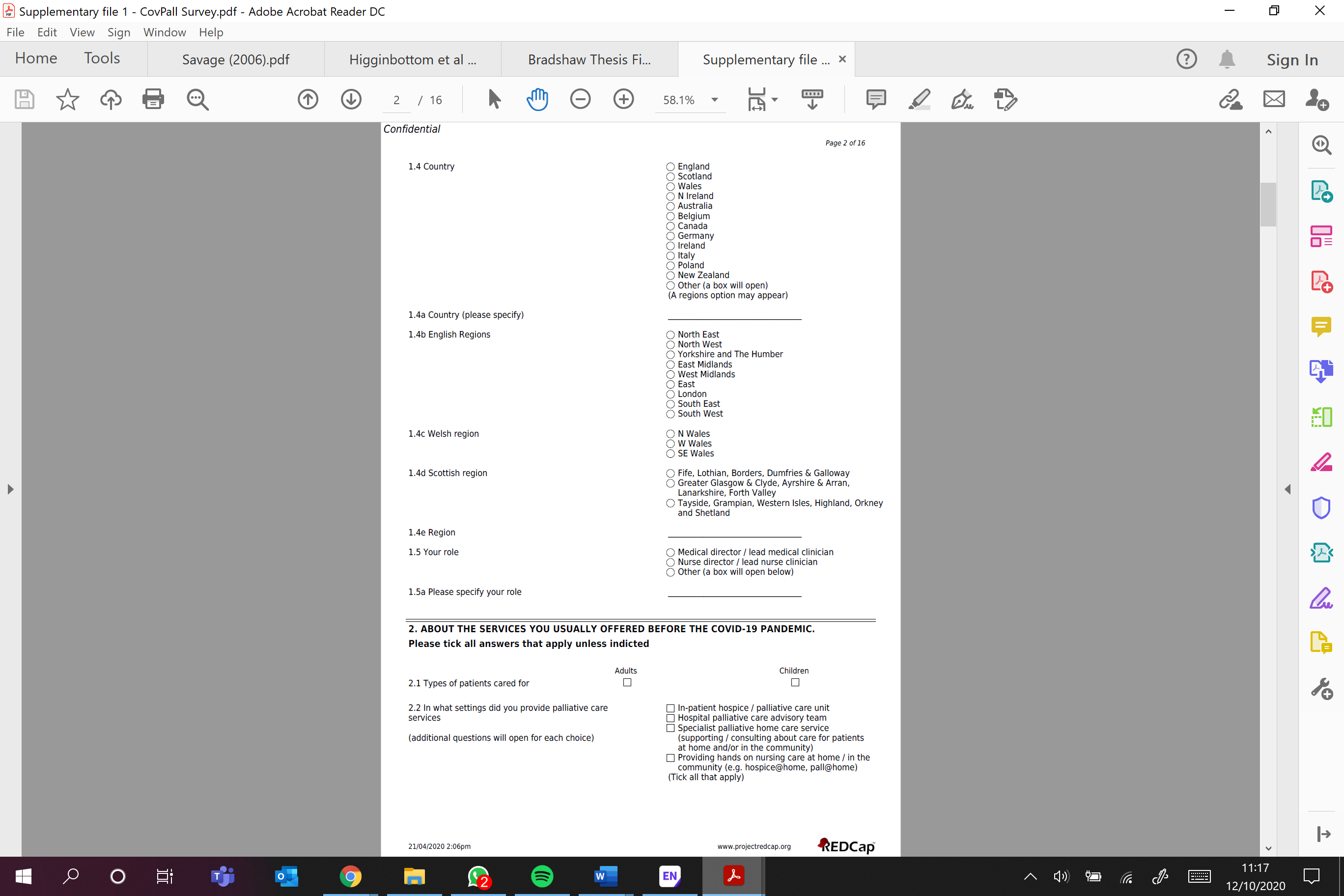

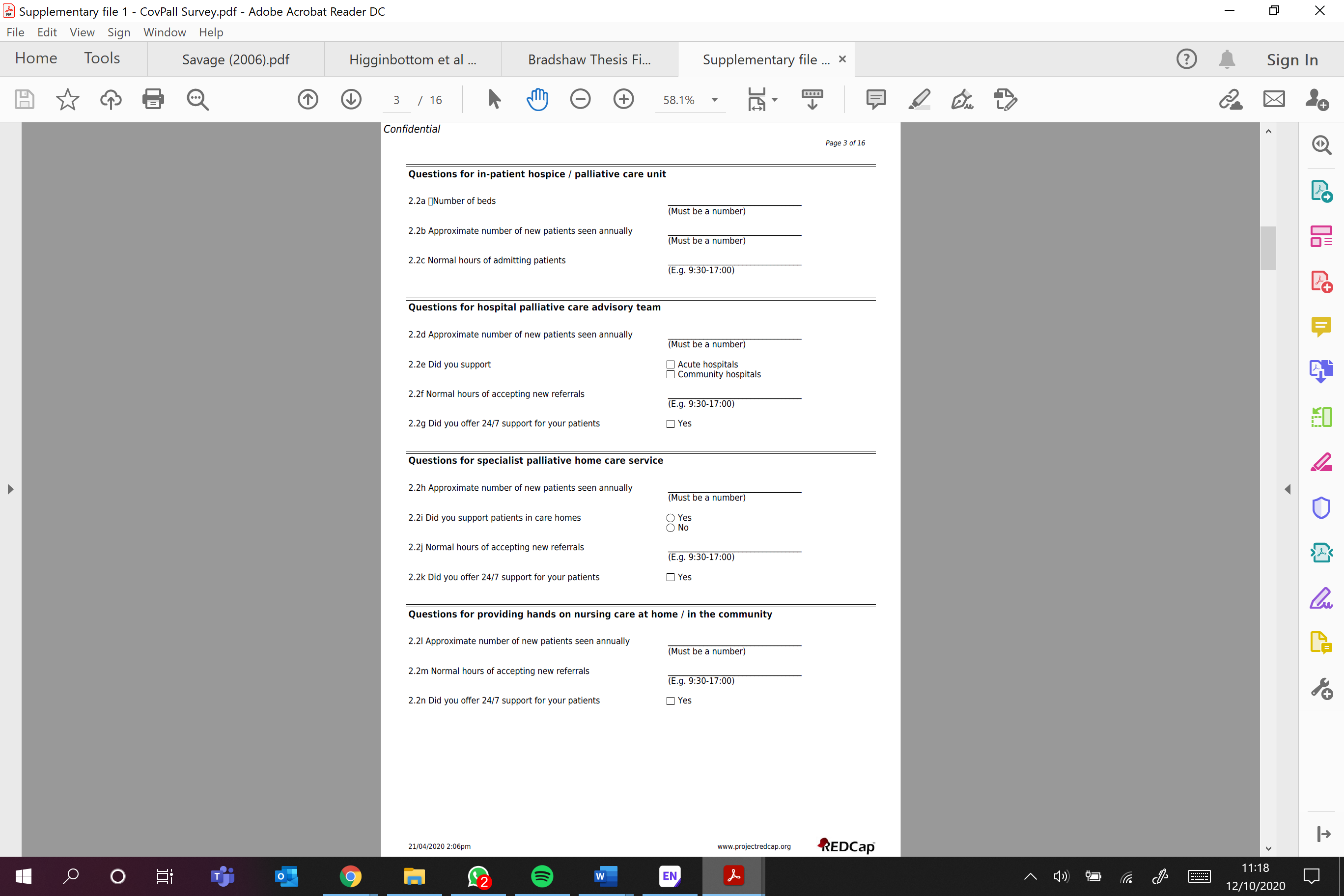

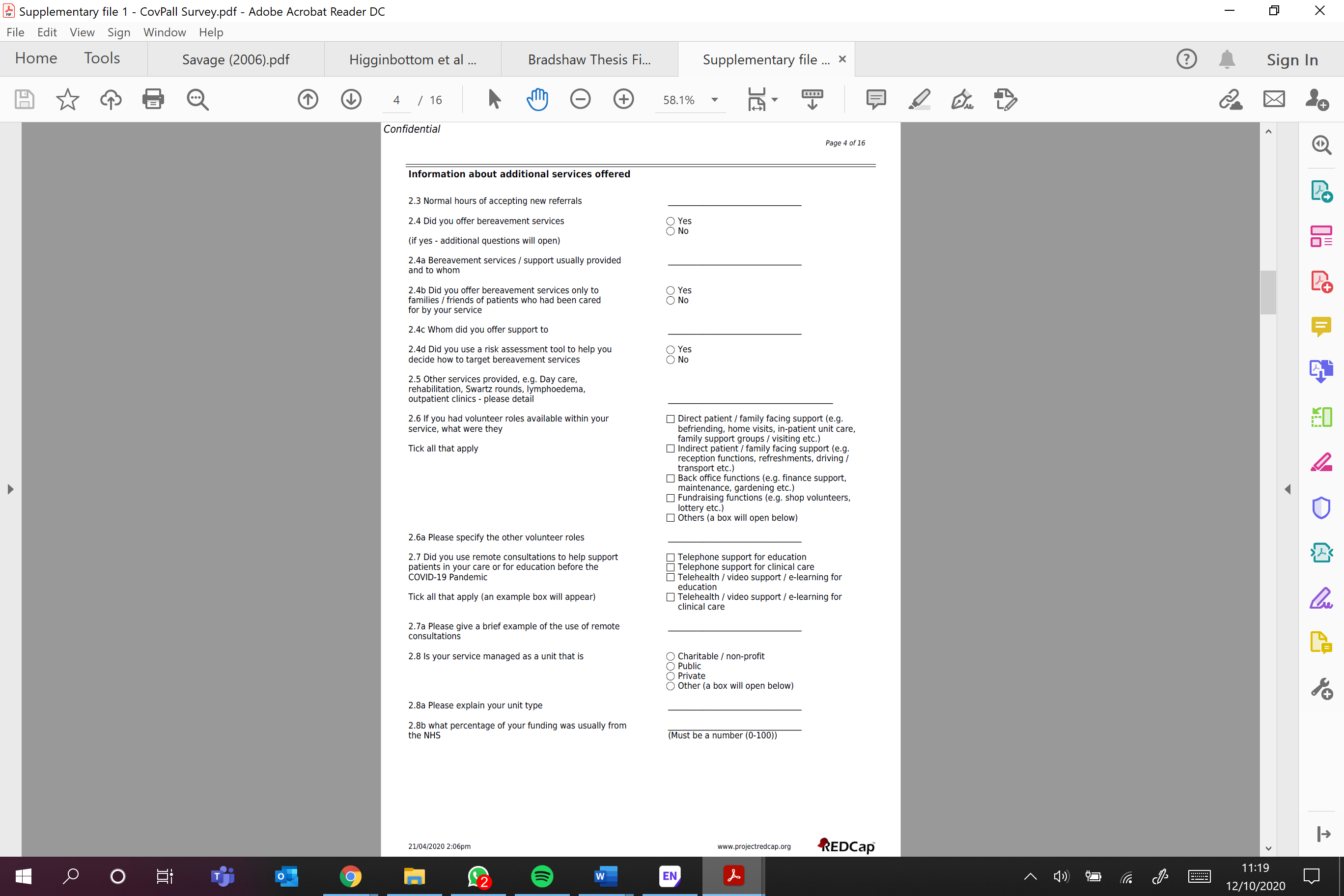

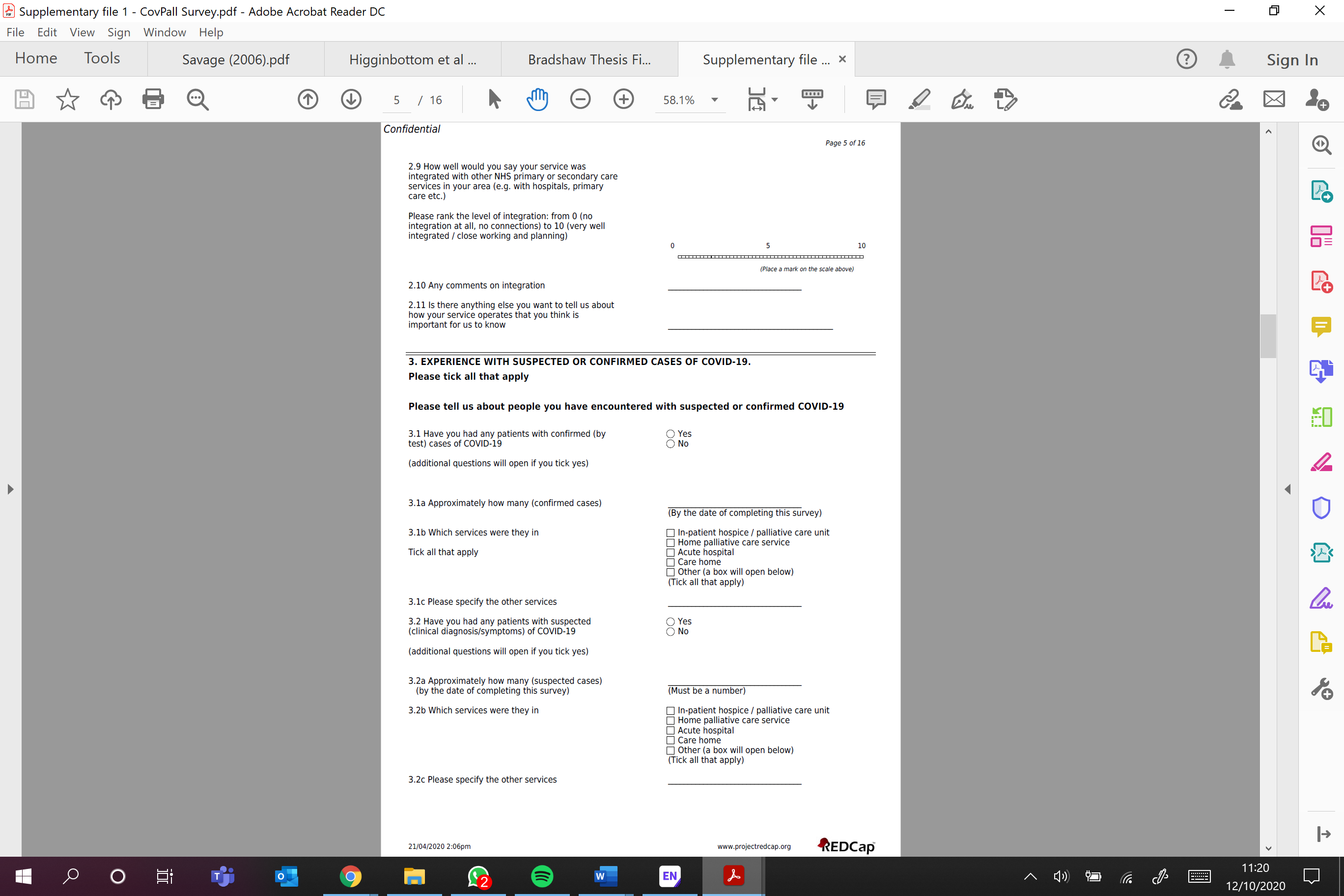

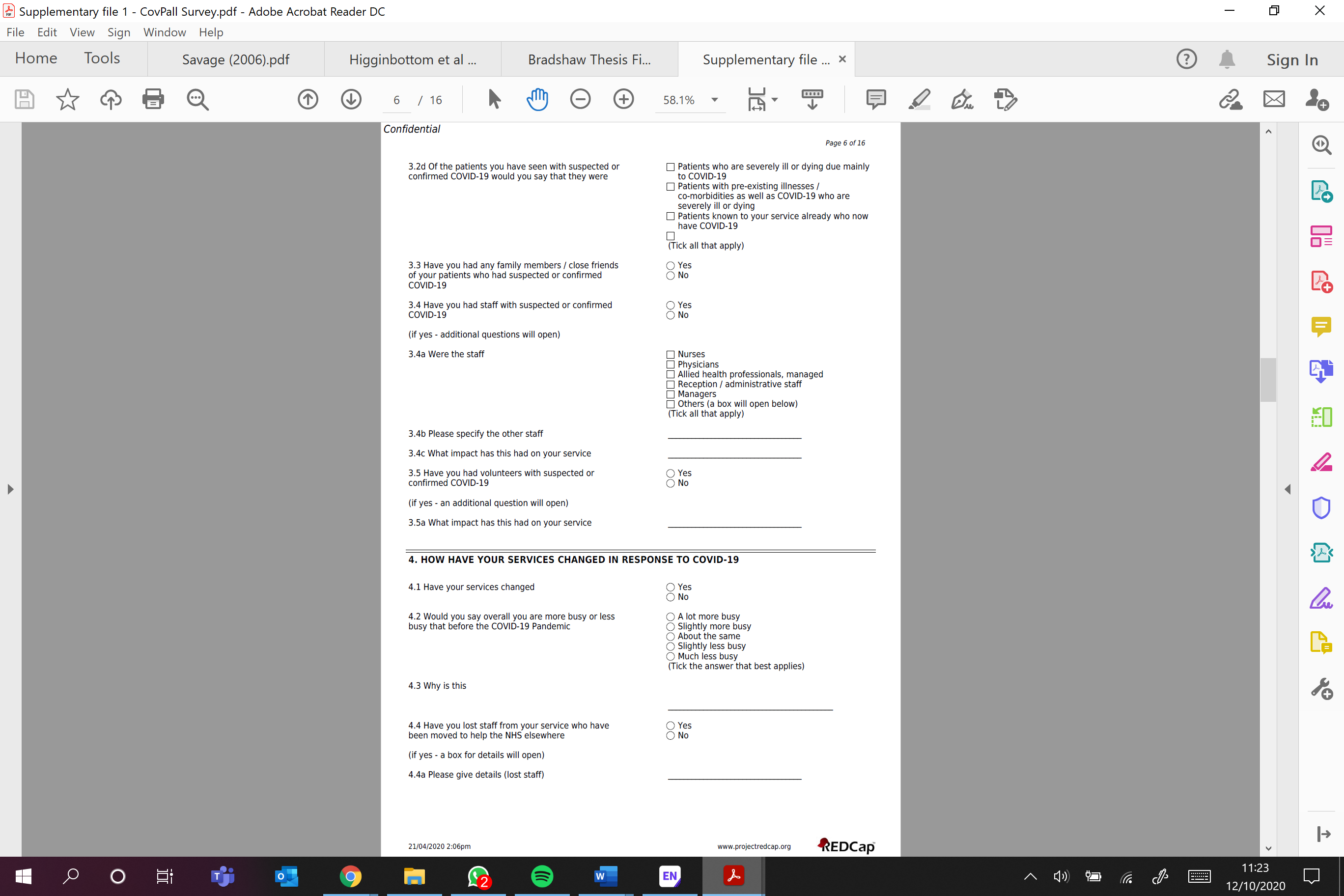

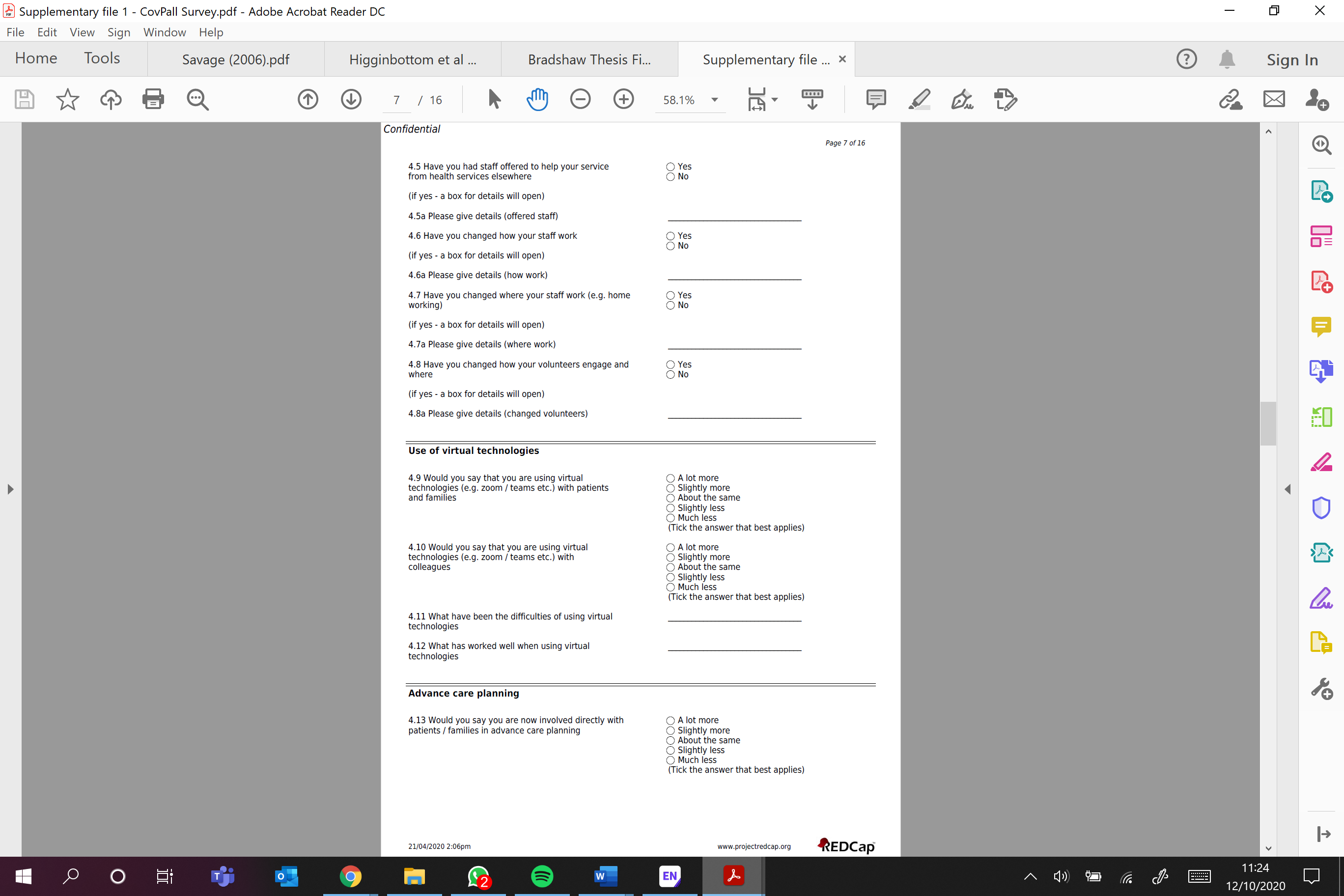

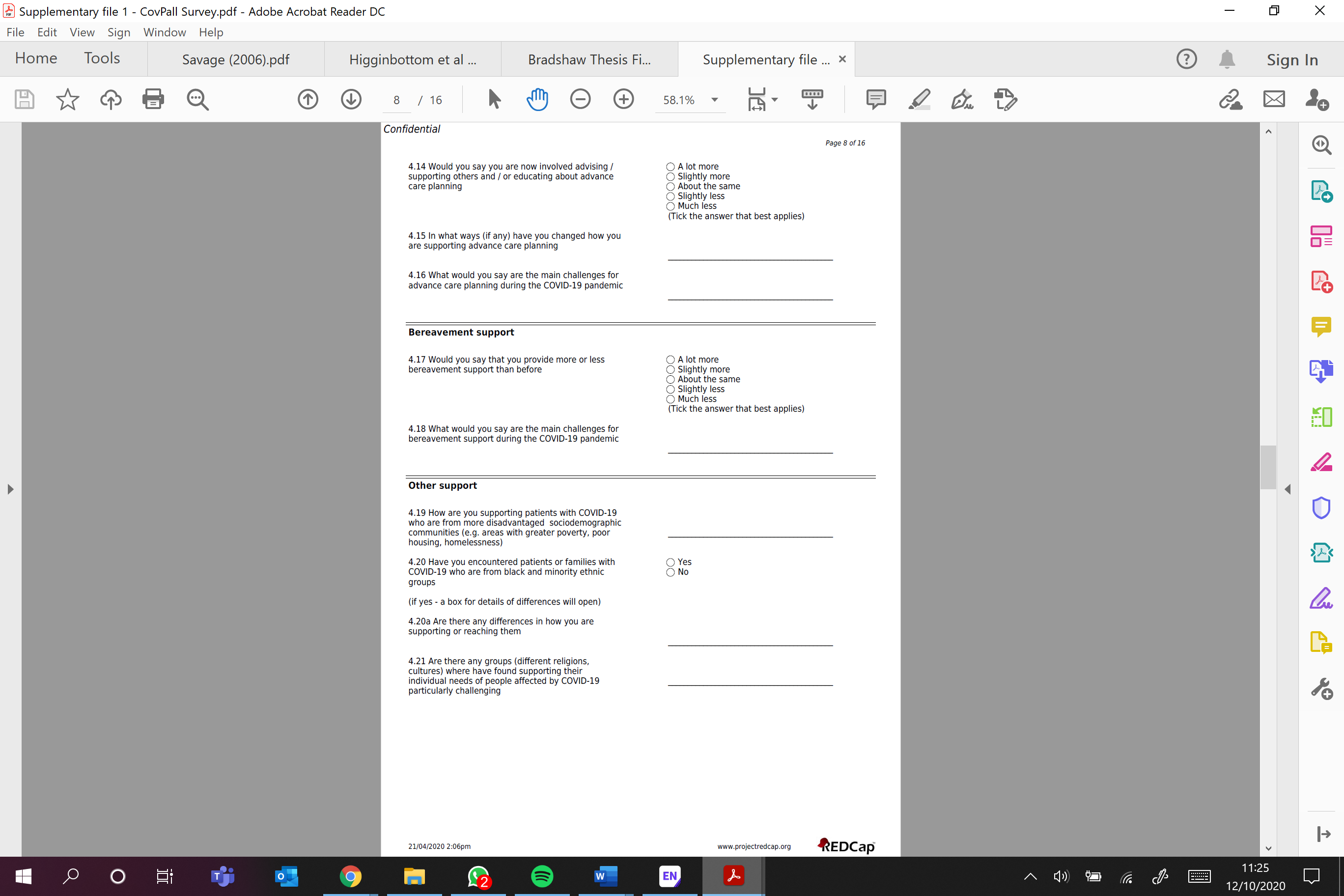

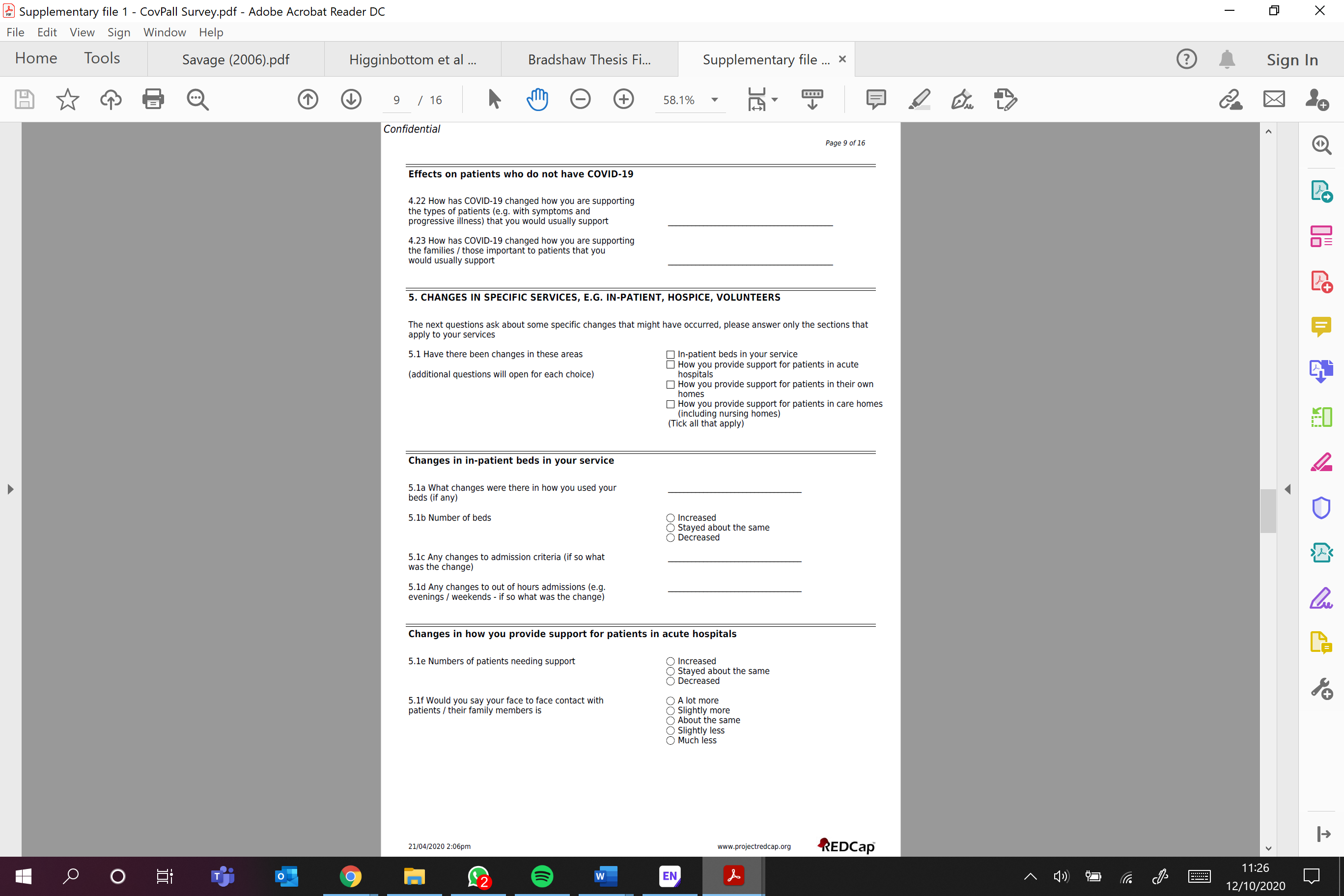

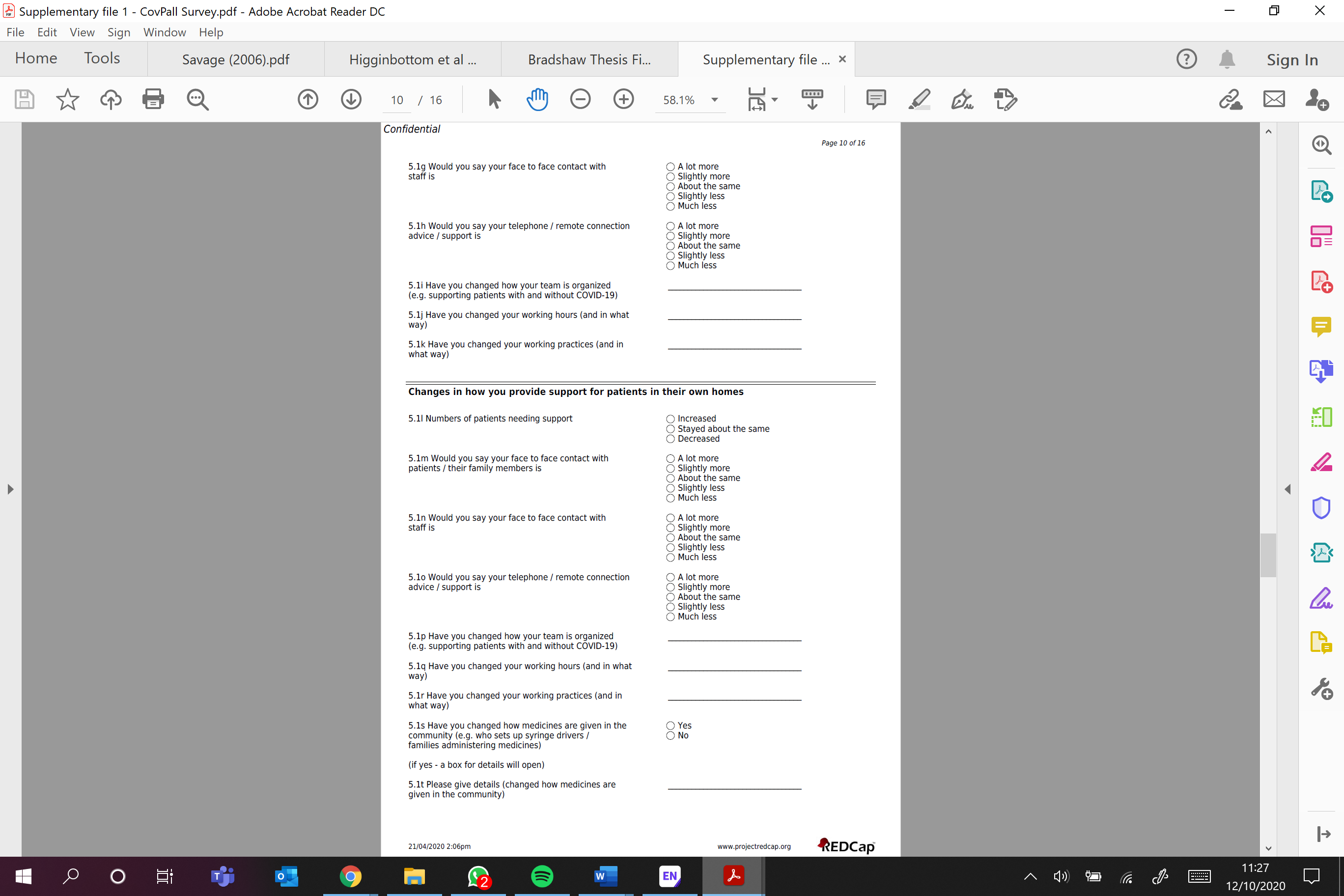

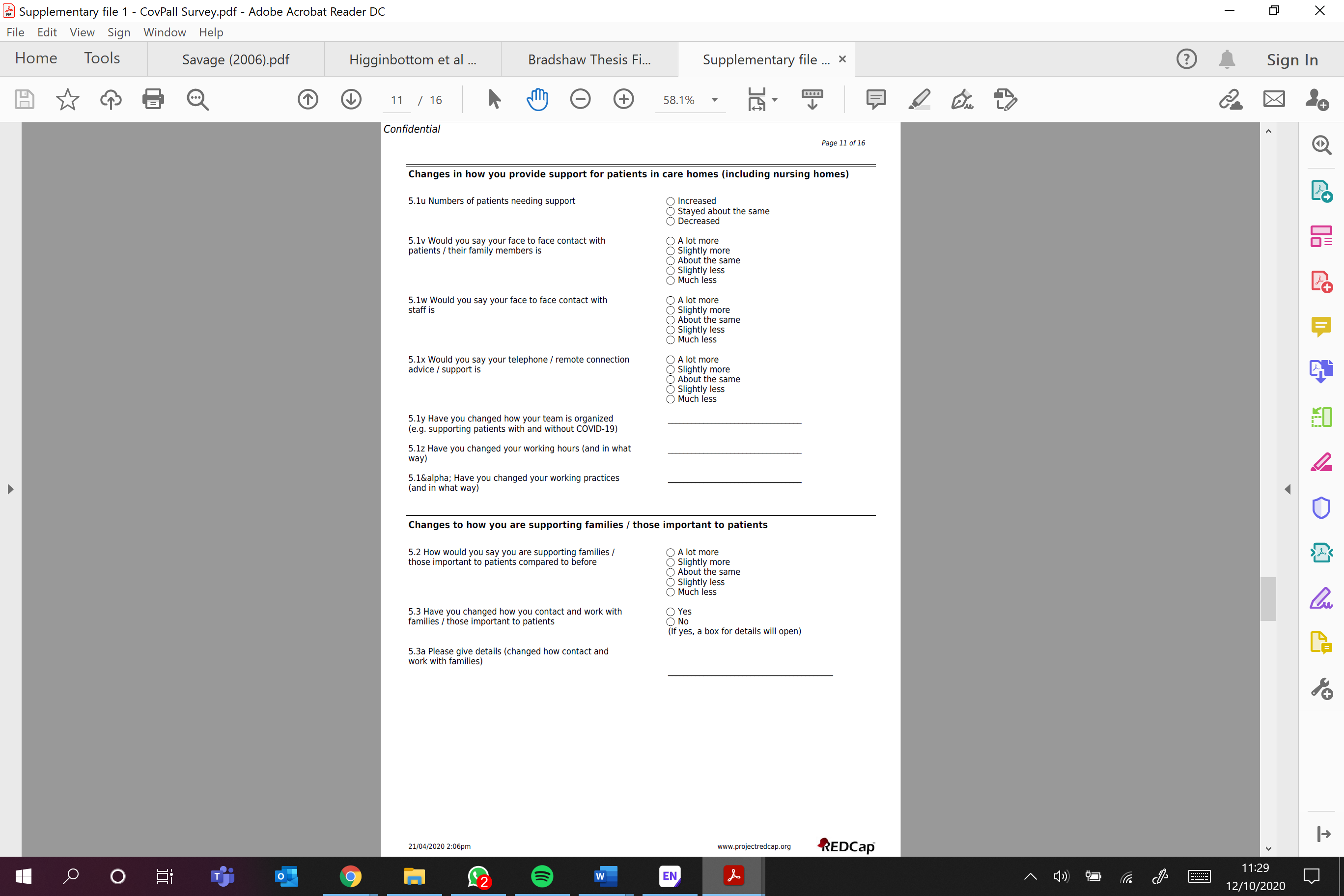

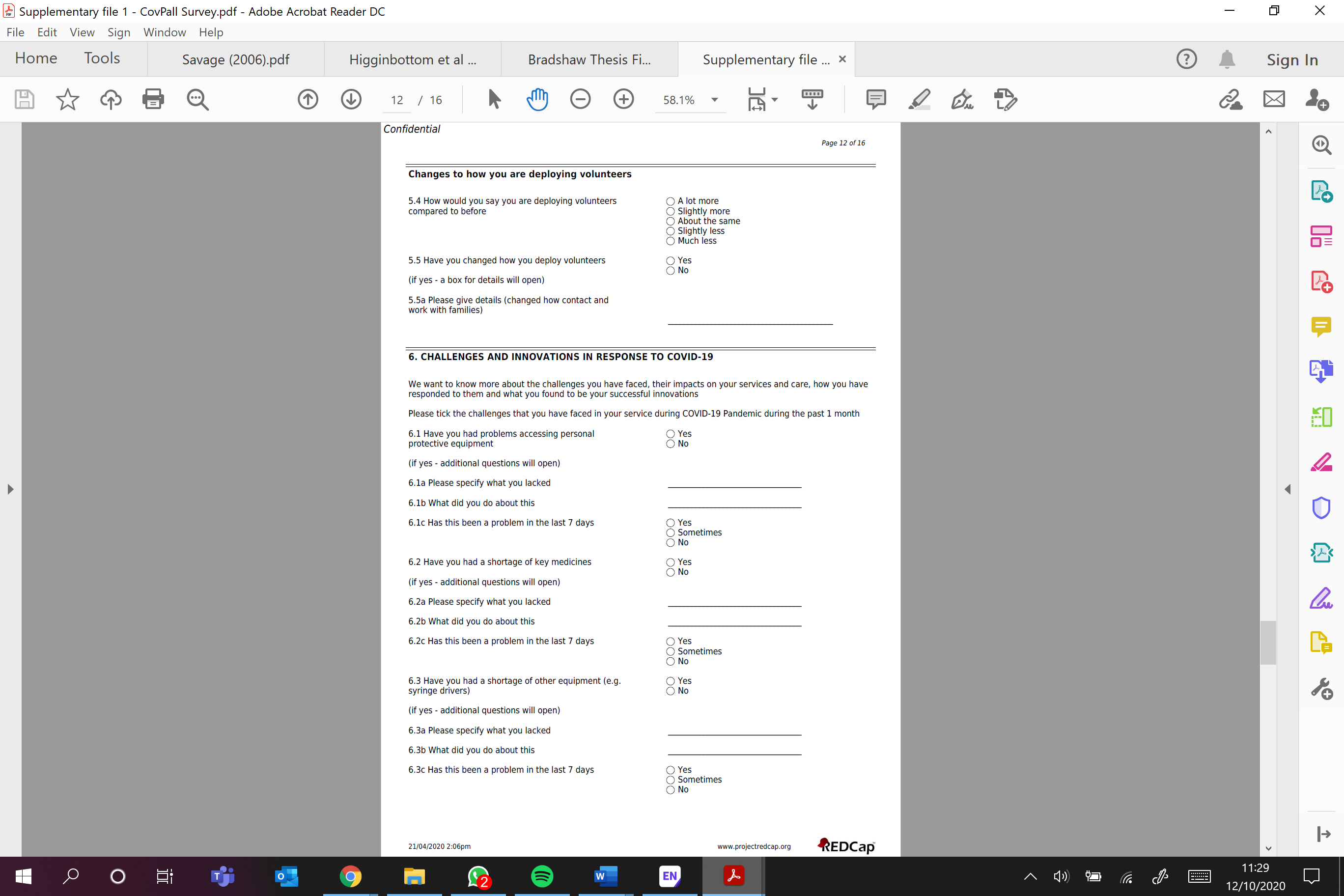

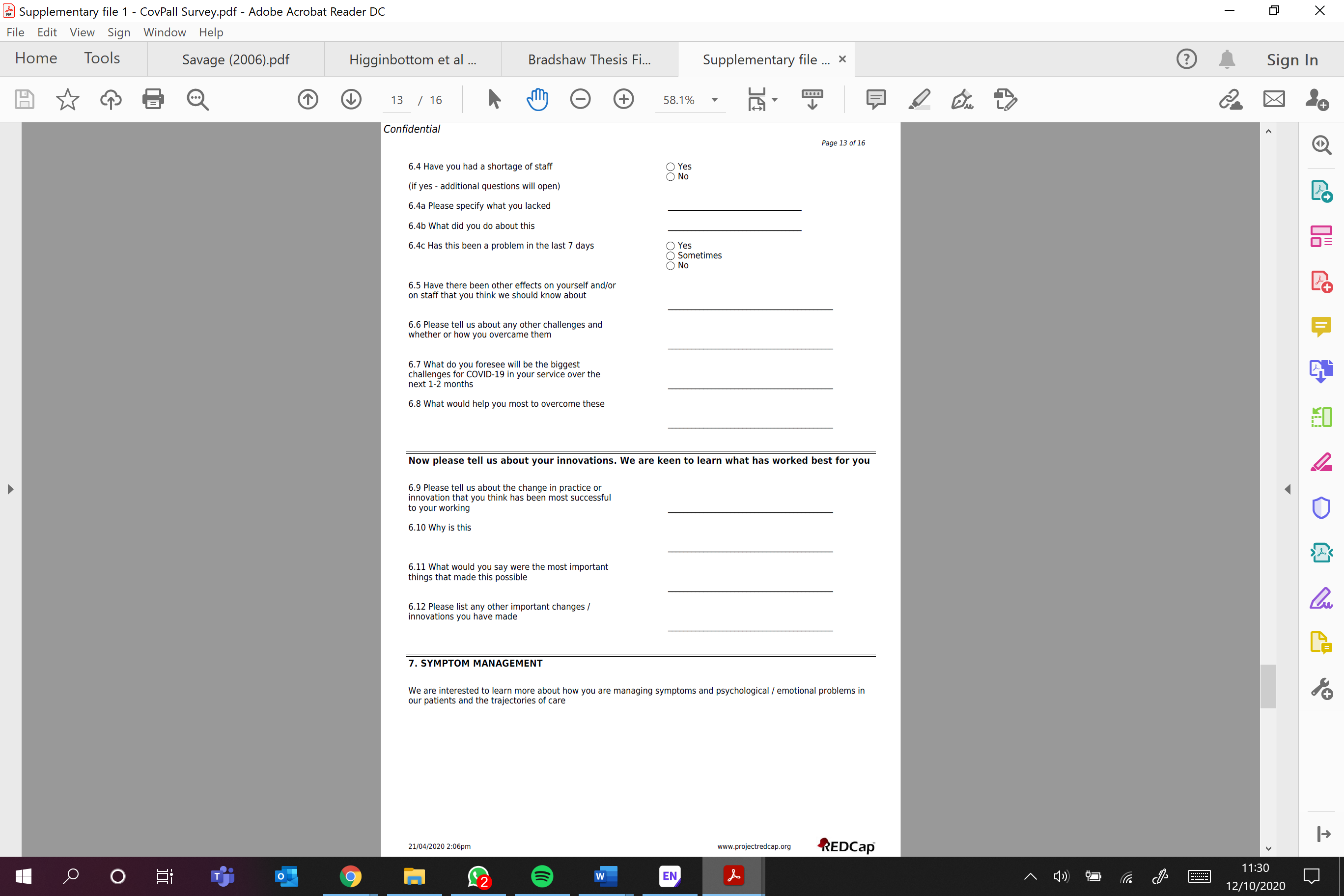

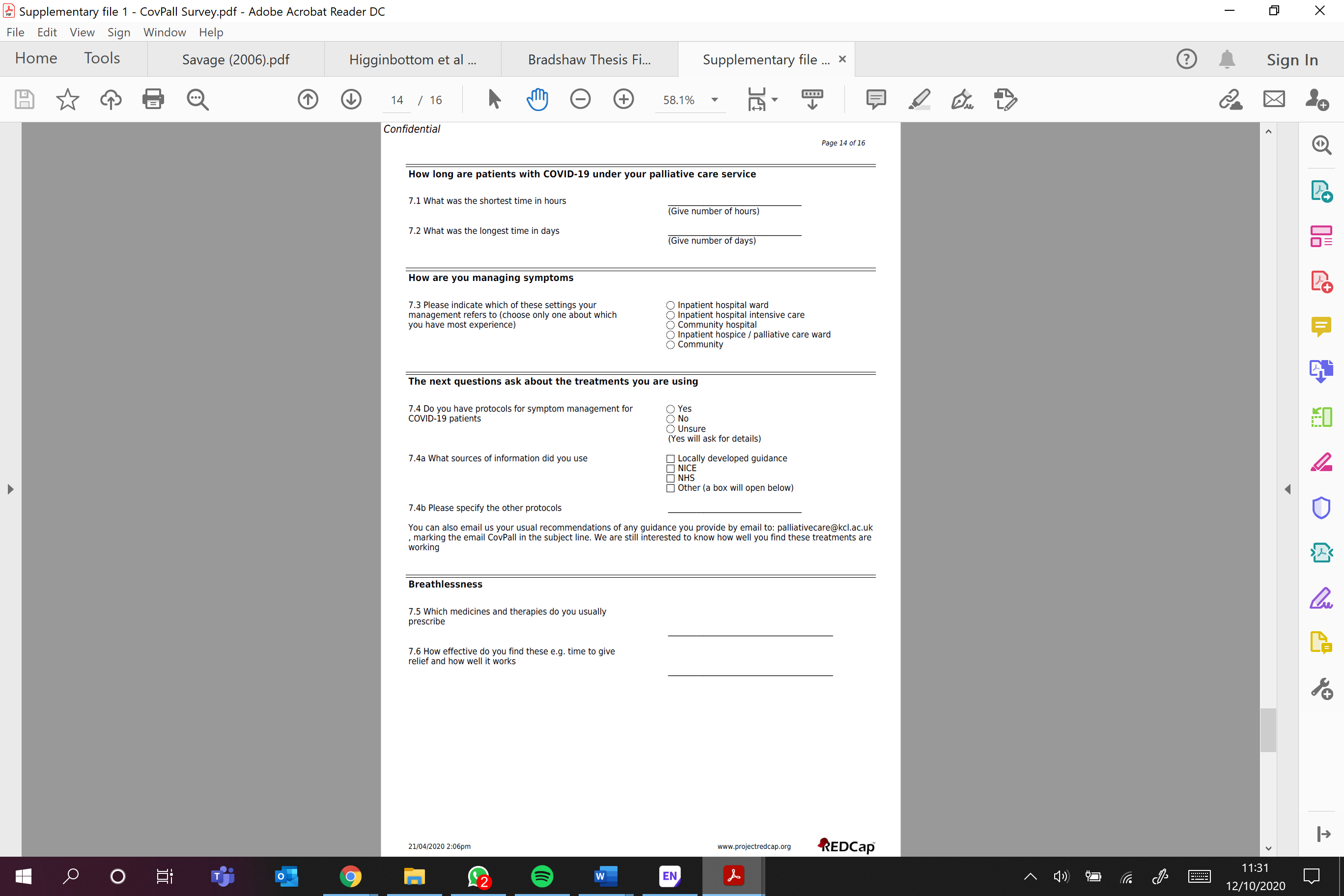

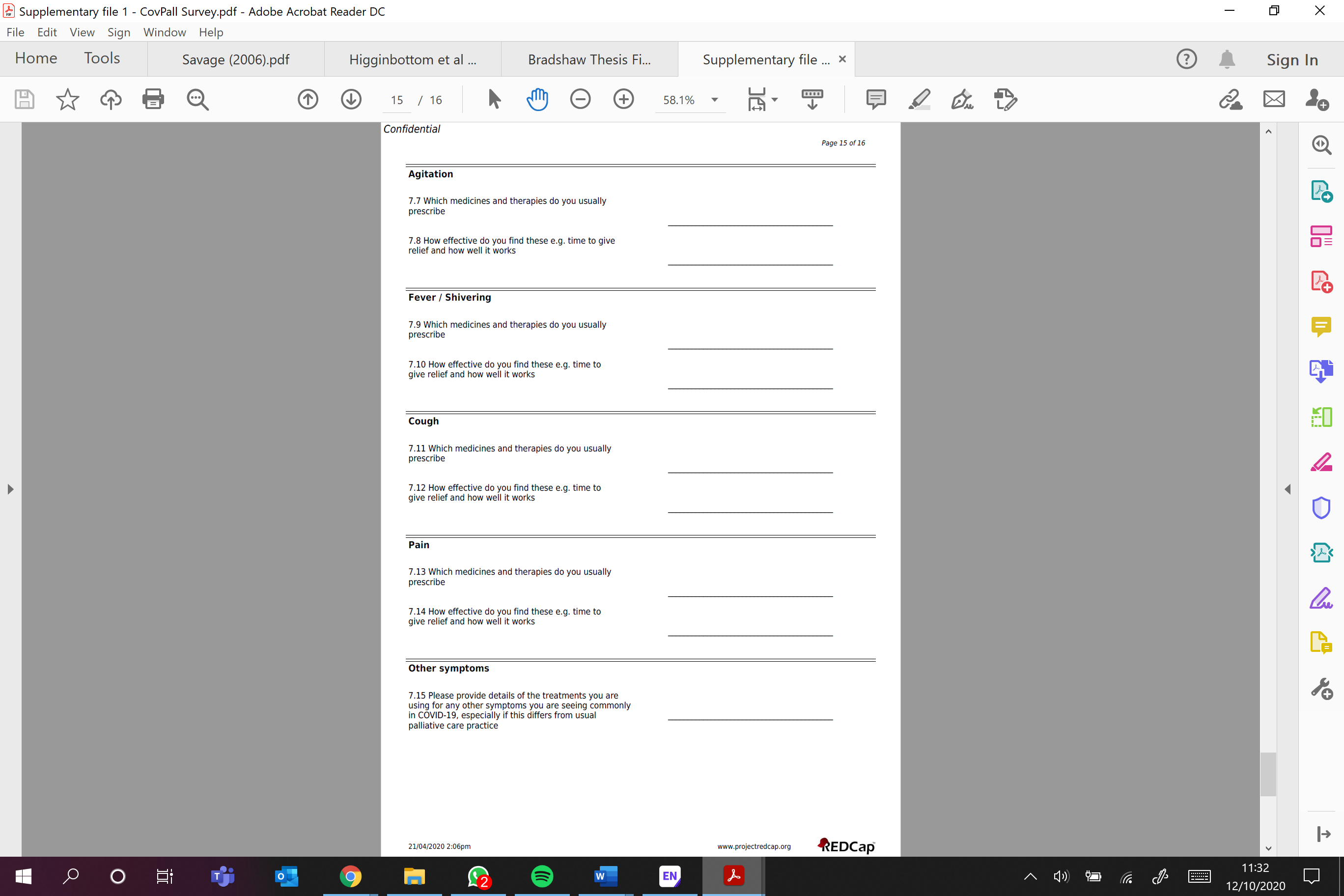

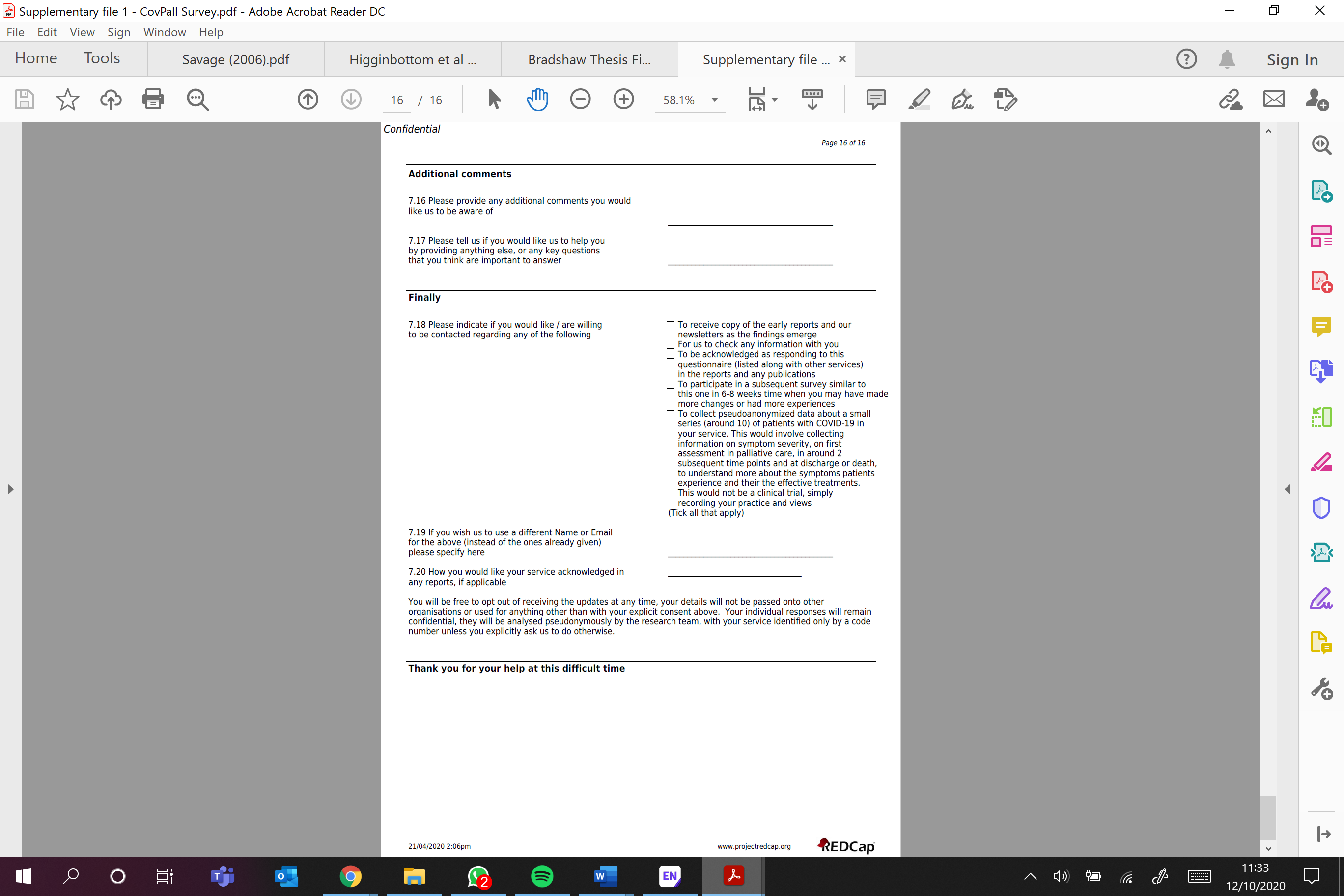
