## Supplementary file 4 for "‘Necessity is the mother of invention’: Specialist palliative care service innovation and practice change in response to COVID-19. Results from a multi-national survey (CovPall)"

STROBE Statement—Checklist of items that should be included in reports of ***cross-sectional studies***

|  | Item No | Recommendation | Page No |
| --- | --- | --- | --- |
| **Title and abstract** | 1 | (*a*) Indicate the study’s design with a commonly used term in the title or the abstract | Yes page 1 |
|  |  | (*b*) Provide in the abstract an informative and balanced summary of what was done and what was found | Yes page 2 |
| Introduction | | | |
| Background/rationale | 2 | Explain the scientific background and rationale for the investigation being reported | Yes pages 4 and 5 |
| Objectives | 3 | State specific objectives, including any prespecified hypotheses | Yes page 5 |
| Methods | | | |
| Study design | 4 | Present key elements of study design early in the paper | Yes page 5 |
| Setting | 5 | Describe the setting, locations, and relevant dates, including periods of recruitment, exposure, follow-up, and data collection | Yes pages 5 and 8 |
| Participants | 6 | (*a*) Give the eligibility criteria, and the sources and methods of selection of participants | Yes page 5 |
| Variables | 7 | Clearly define all outcomes, exposures, predictors, potential confounders, and effect modifiers. Give diagnostic criteria, if applicable | Table one (pages 6 and 7) contains details of the survey questions relevant to this paper |
| Data sources/ measurement | 8* | For each variable of interest, give sources of data and details of methods of assessment (measurement). Describe comparability of assessment methods if there is more than one group | Online survey questions (see supplementary data for full survey questions) |
| Bias | 9 | Describe any efforts to address potential sources of bias | Anonymised data prior to analysis  Page 8 |
| Study size | 10 | Explain how the study size was arrived at | The aim was to reach as many specialist palliative care services as possible via key collaborators. The response rate could not be calculated as the survey denominator was unknown. Pages 5 and 8 |
| Quantitative variables | 11 | Explain how quantitative variables were handled in the analyses. If applicable, describe which groupings were chosen and why | Continuous variables were expressed as means (SD) and medians (IQR), categorical variables as counts and percentages. Page 8 |
| Statistical methods | 12 | (*a*) Describe all statistical methods, including those used to control for confounding | descriptive statistics, frequencies, proportions and means, page 8 |
|  |  | (*b*) Describe any methods used to examine subgroups and interactions | N/A in this paper |
|  |  | (*c*) Explain how missing data were addressed | Missing data were not imputed. Page 8 |
|  |  | (*d*) If applicable, describe analytical methods taking account of sampling strategy | N/A |
|  |  | (*e*) Describe any sensitivity analyses | N/A |
| Results | | | |
| Participants | 13* | (a) Report numbers of individuals at each stage of study—eg numbers potentially eligible, examined for eligibility, confirmed eligible, included in the study, completing follow-up, and analysed | N/A  Number included in the study and analysed page 8 |
|  |  | (b) Give reasons for non-participation at each stage | N/A |
|  |  | (c) Consider use of a flow diagram | N/A |
| Descriptive data | 14* | (a) Give characteristics of study participants (eg demographic, clinical, social) and information on exposures and potential confounders | See table one pages 9-15  N/A |
|  |  | (b) Indicate number of participants with missing data for each variable of interest | See table one pages 9-15 |
| Outcome data | 15* | Report numbers of outcome events or summary measures | N/A |
| Main results | 16 | (*a*) Give unadjusted estimates and, if applicable, confounder-adjusted estimates and their precision (eg, 95% confidence interval). Make clear which confounders were adjusted for and why they were included | N/A |
|  |  | (*b*) Report category boundaries when continuous variables were categorized | N/A |
|  |  | (*c*) If relevant, consider translating estimates of relative risk into absolute risk for a meaningful time period | N/A |
| Other analyses | 17 | Report other analyses done—eg analyses of subgroups and interactions, and sensitivity analyses | See page 8 for how survey free text comments were analysed |
| Discussion | | | |
| Key results | 18 | Summarise key results with reference to study objectives | Yes page 31 |
| Limitations | 19 | Discuss limitations of the study, taking into account sources of potential bias or imprecision. Discuss both direction and magnitude of any potential bias | Yes pages 32-33 |
| Interpretation | 20 | Give a cautious overall interpretation of results considering objectives, limitations, multiplicity of analyses, results from similar studies, and other relevant evidence | Yes pages 31-33 |
| Generalisability | 21 | Discuss the generalisability (external validity) of the study results | The findings of this large multi- national online survey have been discussed in the context of specialist palliative care nationally and internationally.  Pages 31-33 |
| Other information | | | |
| Funding | 22 | Give the source of funding and the role of the funders for the present study and, if applicable, for the original study on which the present article is based | Yes page 34 |

*Give information separately for exposed and unexposed groups.

**Note:** An Explanation and Elaboration article discusses each checklist item and gives methodological background and published examples of transparent reporting. The STROBE checklist is best used in conjunction with this article (freely available on the Web sites of PLoS Medicine at http://www.plosmedicine.org/, Annals of Internal Medicine at http://www.annals.org/, and Epidemiology at http://www.epidem.com/). Information on the STROBE Initiative is available at www.strobe-statement.org.
